# Clinical Trial of a Probiotic and Herbal Supplement for Lung and Gut Health

**DOI:** 10.1101/2023.01.24.23284954

**Authors:** Nancy M. Wenger, Luhua Qiao, Teodora Nicola, Zoha Nizami, Isaac Martin, Brian A. Halloran, Kosuke Tanaka, Michael Evans, Xin Xu, Timothy G. Dinan, Charles Kakilla, Gillian DunnGalvin, Namasivayam Ambalavanan, Kent A. Willis, Amit Gaggar, Charitharth Vivek Lal

## Abstract

**Rationale:** Dysbiosis of the gut microbiome may augment lung disease via the gut-lung axis. Proteobacteria may increase MMP-9 release and contribute to tissue proteolysis followed by neutrophil recruitment, lung tissue injury, and perpetuation of chronic lung disease.

**Trial Design:** We sought to determine if a *Lactobacillus* probiotic and herbal blend was safe and well-tolerated in healthy volunteers and asthmatic patients.

**Methods:** We conducted a 1-month randomized, open-label clinical trial in Cork, Ireland with healthy and asthmatic patients who took the blend twice a day. The primary endpoint was safety with secondary endpoints including quality of life, lung function, gut microbiome ecology, and inflammatory biomarkers.

**Results:** All subjects tolerated the blend without adverse events. Asthmatic subjects who took the blend showed significant improvements in lung function as measured by forced expiratory volume and serum short chain fatty acid levels from baseline to Week 4. The gut microbiome of asthmatic subjects differed significantly from controls, with the most prominent difference in the relative abundance of the proteobacteria *Escherichia coli*. Administration of the probiotic maintained overall microbial community architecture with the only significant difference being an increase in absolute abundance of the probiotic strains measured by strain-specific PCR.

**Conclusions:** This study supports the safety and efficacy potential of a *Lactobacillus* probiotic plus herbal blend to act on the gut-lung axis.

ClinicalTrials.gov Identifier: NCT05173168

## Introduction

The gut-lung axis serves as a powerful means of communication between the microbiome of the gut and the inflammatory and immune microenvironment of the lungs. Patients with respiratory diseases often have gastrointestinal symptoms and show distinct imbalanced gut microbiomes compared to healthy individuals (1-5). Inhaled exposure to environmental toxins such as cigarette smoke and pollution can decrease bacterial diversity and increase inflammation in the gut (6, 7). As such, the state of the intestinal microbiota is deeply connected to lung health and vice versa.

Supplementation with probiotic lactic-acid-producing bacteria has been widely studied. Live strains from the genus *Lactobacillus* have been individually clinically studied for supporting gut and lung health in improving and preventing infections (8, 9). Commensal species *L. rhamnosus, L. plantarum*, and *L. acidophilus* support maintaining proper uptake of short chain fatty acids (SCFAs) in both healthy and diseased populations (10-12). SCFAs and other metabolites produced by commensal bacteria travel through systemic circulation and mediate inflammatory and immune responses in distal organs (13). In a double-blind, randomized controlled trial of asthmatic patients, a *Lactobacillus* blend taken once a day for 8 weeks showed immunomodulatory effects via improvement in Th2 cells-associated IL-4 and lung function via forced expiratory volume and forced vital capacity (14). Adequate SCFA production is also associated with reduced allergic airway inflammation (15, 16).

Bioactive compounds in herbal extracts can further support the anti-inflammatory action of live probiotic strains. Herbal extracts of holy basil leaf, turmeric root, and vasaka leaf can be taken orally to support antioxidant, antitussive, anti-inflammatory, and bronchodilatory effects in the lungs and systemically (17-19). A blend of live *Lactobacillus* strains with holy basil, turmeric, and vasaka extracts, decreased matrix metalloproteinase 9 (MMP-9) pathway activity, neutrophil recruitment, and pro-inflammatory biomarkers in preclinical *in vitro* and *in vivo* models of lung inflammation (20).

Our group has focused on attenuating inflammation and supporting lung function through the gut-lung axis using oral probiotic supplementation. In this clinical study, we hypothesized that a preclinically studied (20) *Lactobacillus* probiotic and herbal blend would demonstrate safety and preliminary biomarker and clinical improvements in healthy and asthmatic populations in a 1-month course of dosing.

## Results

### Oral supplementation with probiotic and herbal blend is safe for human consumption

Forty total participants were screened to randomize 22 participants with asthma (n=11) or were healthy (n=11) who met study eligibility criteria (Table 1, Figure 1). Smoking status was noted (healthy smokers n=3, asthmatic smokers n=4) because of its well-documented impact on lung health and susceptibility to disease. Participants who met the eligibility criteria and successfully completed the Screening Visit were enrolled in the trial. Subjects took the probiotic blend twice a day for 4 weeks, and assessments were conducted at Baseline and the end of Week 2 and Week 4.

**Table 1.**
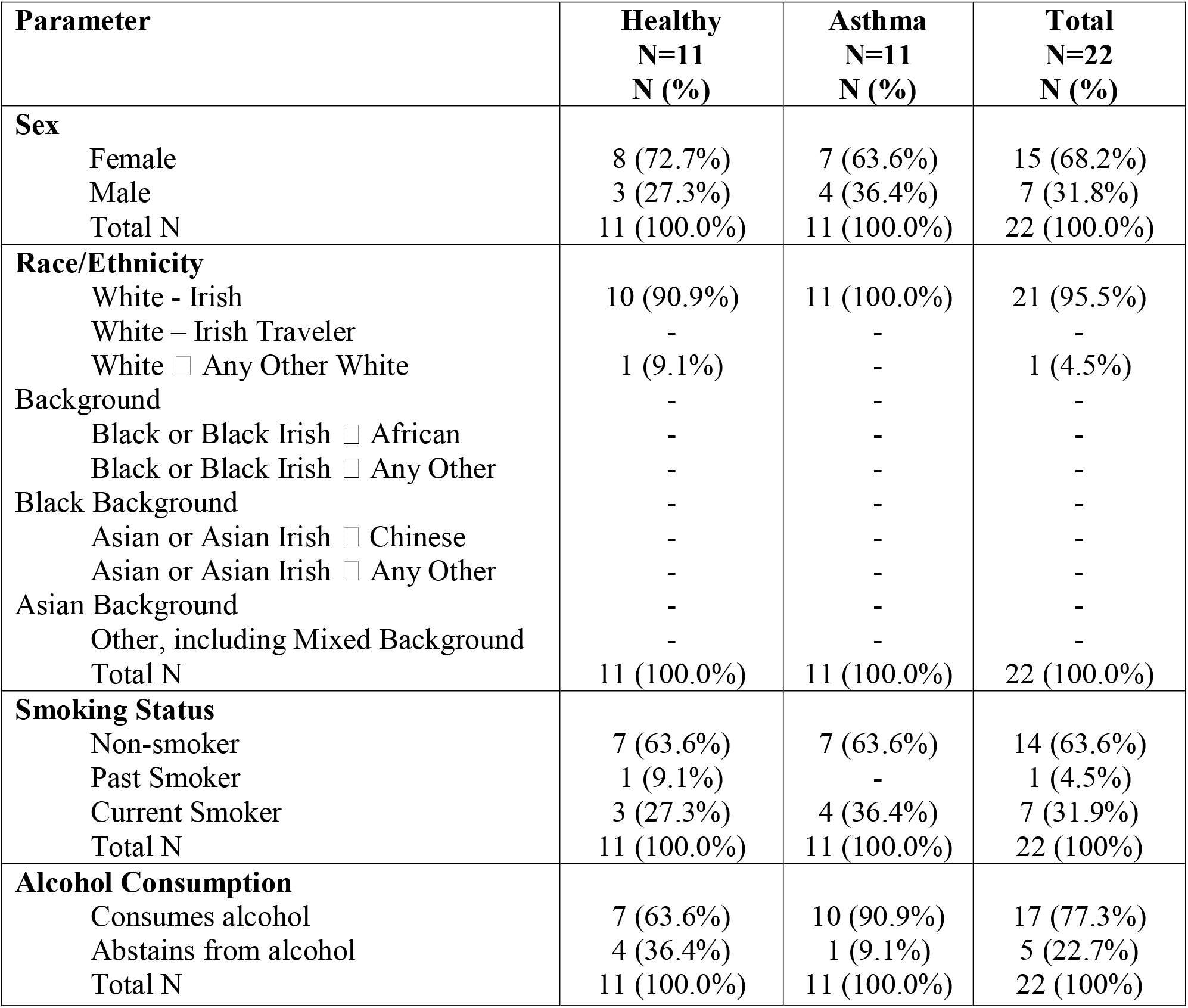
Frequency table of participant demographics collected at baseline in the safety population.

**Figure 1.**
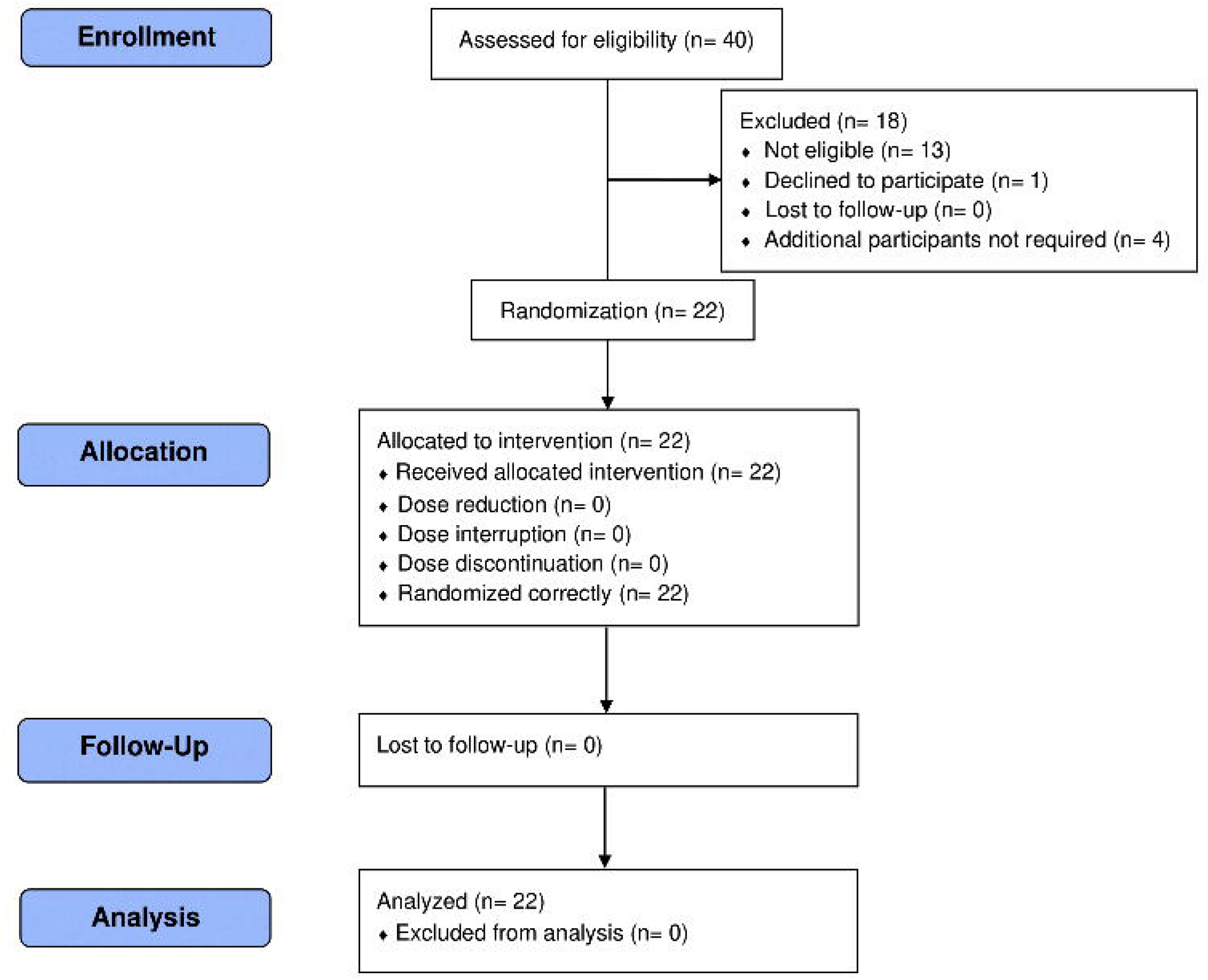
CONSORT flow diagram.

No SAEs or withdrawals due to AEs occurred during this study in the healthy and asthmatic populations, successfully satisfying our primary endpoint (Table 2). There was only one mild gastrointestinal complaint of bloating, which required no action, reported during the trial (Supplemental Table 1). Individual results for vital signs and safety blood parameters were clinically reviewed by the medical doctor and deemed to be safe at all timepoints across all participants (Supplemental Tables 2-3).

**Table 2.** Frequency table for AEs and SAEs by health status and MEDRA SOC in the safety population.

| **Health Status** |  | *N* | % |
| --- | --- | --- | --- |
| Healthy | No SAE Reported | 11 | 100.0 |
| Asthma | No SAE Reported | 11 | 100.0 |
| Healthy | No AE Reported | 6 | 54.5 |
|  | Possibly related AE<br>○ Blood pressure increased<br>○ Blood urea increased | 2 | 18.2 |
|  | Probably related AE<br>○ Abdominal distension | 1 | 9.1 |
|  | Not related AE | 0 | 0.0 |
|  | Unlikely related AE<br>○ Hyperphosphatemia<br>○ Tachycardia | 2 | 18.2 |
|  | Total | 11 | 100.0 |
| Asthma | No AE Reported | 8 | 72.7 |
|  | Possibly related AE | 0 | 0.0 |
|  | Probably related AE | 0 | 0.0 |
|  | Not related AE<br>○ Lower respiratory tract infection | 2 | 18.2 |
|  | Unlikely related AE<br>○ Urinary tract infection | 1 | 9.1 |
|  | Total | 11 | 100.0 |

### Probiotic and herbal blend improves lung function in asthmatic subjects

Spirometry measurements were taken at Baseline and Week 4 to assess changes in lung function. In asthmatic participants, average FEV1% increased significantly (*P*=0.018) and FVC trended up (*P*=0.082) from Baseline to Week 4 (Table 3). The healthy population did not see a change in FEV1% (*P*=0.099) or FVC (*P*=0.387), and neither group had a significant improvement in FEV/FVC ratio (healthy *P*=0.113, asthma *P*=0.284).

**Table 3.** Paired samples test to assess within group change in lung function parameters from Baseline to Week 4 in the healthy (n=11) and asthma (n=11) populations.

|  | Paired Differences |  |  |  |  |  |  |  |
| --- | --- | --- | --- | --- | --- | --- | --- | --- |
|  |  |  |  | 95% Confidence Interval of the Difference |  |  |  | Significance |
|  | Mean | SD | SEM | Lower | Upper | t | df | Two-sided p |
| Healthy<br>FEV1 Average (%) Week 4 – Baseline | -5.000 | 9.110 | 2.747 | -11.120 | 1.120 | -1.820 | 10 | 0.099 |
| Healthy<br>FVC Average (%) Week 4 – Baseline | -2.091 | 7.661 | 2.310 | -7.238 | 3.056 | -0.905 | 10 | 0.387 |
| Healthy<br>FEV1/FVC Week 4 – Baseline | -0.036 | 0.069 | 0.021 | -0.082 | 0.010 | -1.735 | 10 | 0.113 |
| Asthma<br>FEV1 Average (%) Week 4 – Baseline | 8.455 | 9.974 | 3.007 | 1.754 | 15.155 | 2.811 | 10 | 0.018 |
| Asthma<br>FVC Average (%) Week 4 – Baseline | 4.364 | 7.487 | 2.257 | -0.666 | 9.393 | 1.933 | 10 | 0.082 |
| Asthma<br>FEV1/FVC Week 4 – Baseline | 0.033 | 0.098 | 0.030 | -0.032 | 0.099 | 1.132 | 10 | 0.284 |

### Probiotic and herbal blend improves quality of life scores as measured by SGRQ

Participants responded to the Saint George’s Respiratory Questionnaire (SGRQ) at Baseline, Week 2, and Week 4 to evaluate the supplement’s effect on quality of life as measured through symptoms, impact, and total score. Across total score and the subcategories of symptoms, activity, and impact there was no significant change from baseline to Week 4 in either the healthy or asthmatic participants (Table 4). However, impact scores in asthmatic participants trended towards significance (*P*=0.065) and a majority of all study subjects reported a decrease (improvement) in all score categories. Of the participants that smoke, have asthma, or have asthma and smoke, 36% noted an improvement in their overall health, 43% noted less frequent coughing, 43% noted fewer instances of feeling short of breath, and 29% noted fewer cough or breathing-related sleep disturbances.

**Table 4.** Paired samples test to assess within group change in SGRQ total score and subscales from Baseline to Week 4 in the healthy (n=11) and asthma (n=11) populations.

|  | Paired Differences |  |  |  |  |  |  |  |
| --- | --- | --- | --- | --- | --- | --- | --- | --- |
|  |  |  |  | 95% Confidence Interval of the Difference |  |  |  | Significance |
|  | Mean | SD | SEM | Lower | Upper | t | df | Two-sided p |
| Healthy SGRQ Total Score Week 4 – Baseline | -1.432 | 2.924 | 0.882 | -3.396 | 0.532 | -1.624 | 10 | 0.135 |
| Healthy SGRQ Symptom Score Week 4 – Baseline | -5.829 | 10.962 | 3.305 | -13.194 | 1.535 | -1.764 | 10 | 0.108 |
| Healthy SGRQ Activity Score Week 4 – Baseline | -0.753 | 4.789 | 1.444 | -3.970 | 2.465 | -0.521 | 10 | 0.614 |
| Healthy SGRQ Impact Score Week 4 – Baseline | -0.468 | 2.623 | 0.791 | -2.230 | 1.294 | -0.592 | 10 | 0.567 |
| Asthma SGRQ Total Score Week 4 – Baseline | -3.594 | 7.967 | 2.402 | -8.946 | 1.758 | -1.496 | 10 | 0.165 |
| Asthma SGRQ Symptom Score Week 4 – Baseline | -10.012 | 20.628 | 6.220 | -23.870 | 2.846 | -1.610 | 10 | 0.139 |
| Asthma SGRQ Activity Score Week 4 – Baseline | 1.333 | 10.153 | 3.061 | -5.487 | 8.154 | -.436 | 10 | 0.672 |
| Asthma SGRQ Impact Score Week 4 | -4.451 | 7.118 | 2.146 | -9.233 | 0.330 | -2.074 | 10 | 0.065 |
| – Baseline |  |  |  |  |  |  |  |  |

### Majority of participants would recommend probiotic and herbal blend

In an end of product study, 100% of participants who smoke would recommend the probiotic and herbal blend to friends and family. 90% of asthmatic participants and 82% of healthy participants would recommend the same.

### Probiotic and herbal blend improves serum short chain fatty acid levels in asthmatic subjects

As we saw no significant changes from baseline to Visit 4 in the selected pro-inflammatory cytokines, we decided to analyze the serum for short chain fatty acid (SCFA) levels from baseline to Visit 4. Participants with asthma showed significant changes in serum SCFA levels. Propionic acid increased across all asthmatics (Figure 2A), propionic acid and isovaleric acid increased significantly in asthmatic non-smokers (Figure 2B), and acetic acid and butyric acid significantly increased in subjects with asthma who smoked (Figure 2C). Across all groups, isobutyric acid and valeric acid did not show a significant change, and hexanoic acid levels were too low for detection.

**Figure 2.**
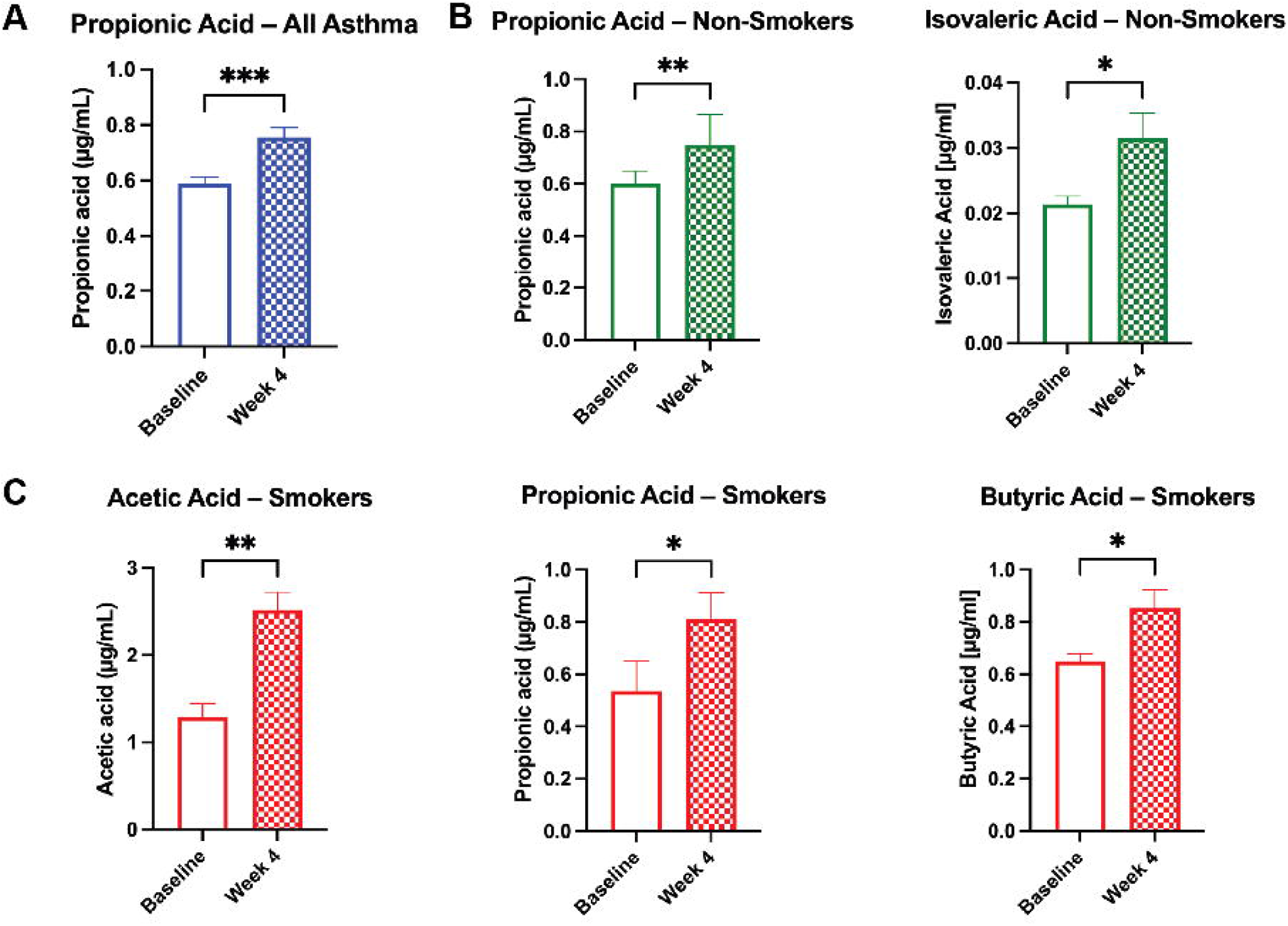
Probiotic and herbal blend improves serum SCFA levels in the asthmatic population. Significant increases in serum short chain fatty acid (SCFA) levels as broken down by **A)** all asthmatic participants, **B)** asthmatic non-smokers, and **C)** asthmatic smokers. \**P*<0.05, \*\**P*<0.01, \*\*\**P*<0.001.

### Gut microbiome analyses

To examine the effects of the probiotic on the intestinal microbiome for potentially adverse signals, we collected stool samples at baseline and Week 4 and analyzed for participants’ gut microbiome signatures. The overall alpha (intra-sample) and beta (inter-sample) diversity was not significantly altered between baseline and after 4 weeks of probiotic administration (Shannon Diversity, *P*=0.644, T-test; Chao1 Richness Index, *P*=0.665, T-test, Figure 3A. P=1, R2=0.007, PERMANOVA; P=0.279, PERMDISP, Figure 3B), with only modest differences in colonization at the genus level (Supplemental Table 4). However, the patient’s asthma status did associate with differences in alpha and beta diversity (Chao1, *P*=0.033 Figure 3C, and *P*=0.017, R2 = 0.0464, PERMANOVA; P=0.52805, PERMDISP, Figure 3D). The most prominent differences at the genus level were higher relative abundance of *E. coli* (Log_2_FC 26.6), *Bacteroidetes dorei* (Log_2_FC 24.2), and *B. ovatus* (Log_2_FC 21.7) (Supplemental Table 5).

**Figure 3.**
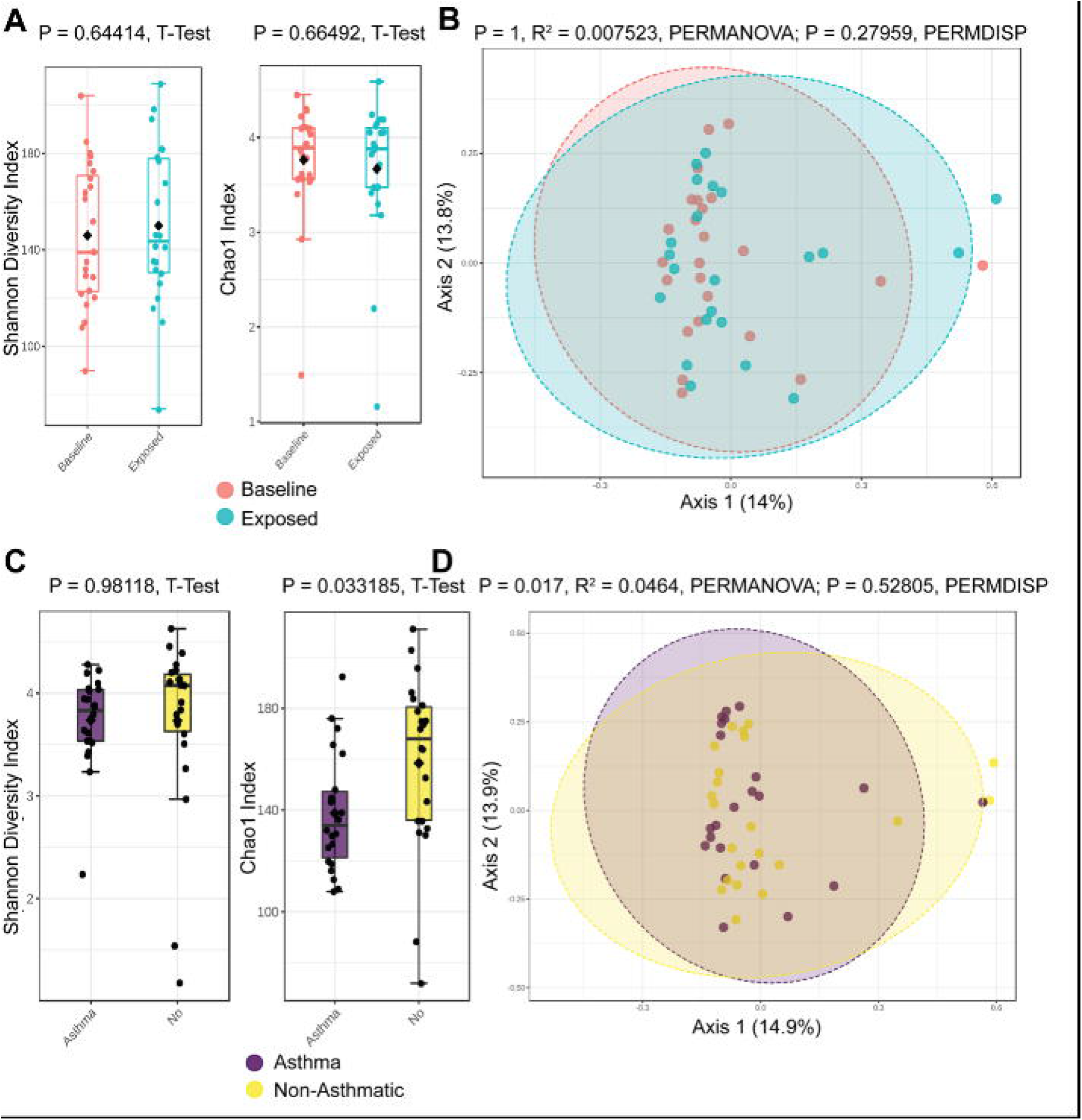
Global microbiome community composition not disrupted by probiotic administration, but asthmatics have a different gut microbiome. (A) Alpha diversity is unaltered by probiotic administration. B) Beta diversity remains similar following probiotic administration. Asthmatic patients have different alpha diversity (C) and beta diversity (D).

### Probiotic *Lactobacillus* strains detected in stool

On a subset of 16 samples with the highest DNA concentrations remaining after MiSeq, we performed qPCR using primers validated for strain-specificity. Most samples were not colonized with *L. plantarum* at baseline (19%), but this colonization was augmented in most samples by administration of the probiotic and herbal blend (81%, chi^2^, 12.5, *P*=0.0004). *L. acidophilus* colonization was more common at baseline (37%) and was detected in most of the post-exposure samples (62%, chi^2^ 2, *P*=0.157). Similarly, for *L. rhamnosus*, colonization was detected in most individuals before and after probiotic administration (62% versus 81%, chi^2^, 1.39, *P*=0.238). In healthy participants, the concentration of *L. plantarum* increased significantly and *L. acidophilus* and *L. rhamnosus* trended upward from baseline to Week 4 after taking the probiotic and herbal blend (Figure 4A). Percent densitometry of *L. plantarum* and *L. acidophilus* increased from baseline to Week 4 with *L. rhamnosus* trending up (Figure 4B). In asthmatic participants, the concentration of all three *Lactobacillus* strains increased significantly from baseline to Week 4 (Figure 4C). Percent densitometry of *L. plantarum* and *L. acidophilus* increased from baseline to Week 4 and *L. rhamnosus* trended upward (Figure 4D).

**Figure 4.**
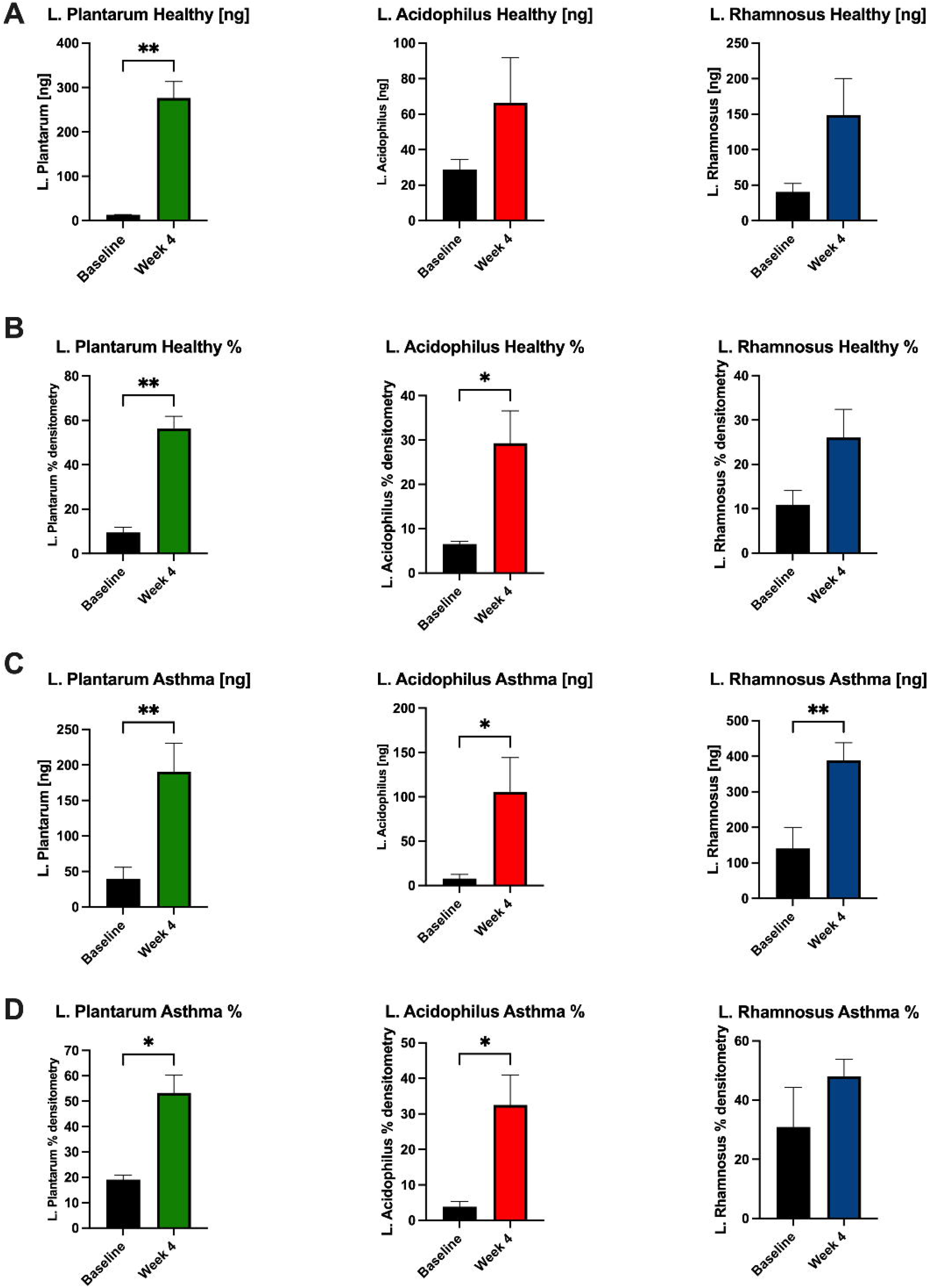
Abundance of probiotic *Lactobacillus* strains increases in stool of participants. In healthy participants, the concentration [ng] of **A)** *L. plantarum* increased significantly and *L. acidophilus* and *L. rhamnosus* trended upward from baseline to Week 4 after supplementation. **B)** % densitometry of *L. plantarum* and *L. acidophilus* increased significantly and *L. rhamnosus* trended upward from baseline to Week 4. In asthmatic participants, the concentration [ng] of **C)** all three *Lactobacillus* strains increased significantly from baseline to Week 4 after supplementation. **D)** % densitometry of *L. plantarum* and *L. acidophilus* increased significantly and *L. rhamnosus* trended upward from baseline to Week 4. \**P*<0.05, \*\**P*<0.01

## Discussion

In our 1-month clinical trial dosing a probiotic and herbal blend to healthy volunteers and asthmatic participants, we found that the supplement was safe and well-tolerated among all subjects, fulfilling our primary endpoint. This was anticipated because prior to this clinical trial, all three probiotic strains have been independently clinically validated and the herbs have a long history of use in preclinical research and traditional medicine (8, 9, 21-24).

A majority of the participants did show a modest increase in quality-of-life scores, although the changes were not statistically significant in the healthy or asthmatic participants. In the healthy participants, their SGRQ scores were generally good at baseline which meant that any change was incremental. Even the asthmatic population that was more likely to start from a lower baseline (higher score) did not show a significant change. A longer trial would be required to allow for enough time to capture changes in perceived symptoms as a result of taking the blend.

SCFA levels were not significantly changed in the healthy population from baseline to Visit 4. However, SCFA levels were significantly upregulated in the serum of the asthmatic participants, suggesting a pre-existing deficiency that was addressed through supplementation. SCFAs are known to affect immune cell function and a variety of inflammatory pathways including TNF-α, IL-2, IL-6, and IL-10 (25). We observed distinct increases in serum SCFAs acetic acid, propionic acid, and butyric acid in subjects with asthma who smoked, and propionic acid and isovaleric acid in subjects with asthma who did not smoke. Gut dysbiosis diminishes metabolism of anti-inflammatory SCFAs, impairing the body’s ability to regulate systemic inflammation and exacerbating allergic lung inflammation (15, 16, 26, 27).

An interesting finding was the significant improvement in lung function in the asthmatic group as measured by FEV1%. This change may have been due to the significant uptick in serum SCFAs which traveled through systemic circulation and reduced inflammation in the lungs. Previous studies have shown an association between *Lactobacillus* administration and immune regulation of Th1/Th2 response, reducing allergic inflammation characteristic of asthma (14, 28). The effect of the herbal extracts can also not be discounted, as turmeric, holy basil, and vasaka have antioxidant and anti-inflammatory properties (18, 29). Although FVC values were not statistically significant, a longer study with more subjects may reveal a larger impact over time.

As anticipated, the probiotic induced no significant alterations in the global gut microbiome community composition of the human participants (30). This agrees with prior literature that demonstrates a limited long-term alteration in the gut microbiota from probiotics in healthy adults, that stems from colonization resistance from the microbiota and host factors that perform ecology maintenance (30-34). We speculate that the ASV aligned to *Lactobacillus murinius* is a miss-identified detection of our probiotic stains, an assertion that is supported by our detection and identification of the probiotic strains by qPCR and the rarity of this strain as a human commensal. Alternatively, prior studies of probiotic administration have suggested that the administration of a *Lactobacillus* probiotic can alter the prevalence of other *Lactobacillus* strains without altering overall community structure (30). We verified by strain-specific PCR that the three *Lactobacillus* strains contained in the probiotic blend increased from baseline to Week 4, of note significantly in the asthmatic population.

We identified robust differences in the microbiota of asthmatic patients in our cohort, confirming prior observations that community composition was associated with altered microbiota (35, 36). Specifically, an increase in proteobacteria *E. coli* was particularly notable, since proteobacteria have been linked with increased risk of respiratory disease (37). These differences are interesting and deserving of further validation in a larger cohort of asthmatic individuals.

In summary, the *Lactobacillus* probiotic and herbal blend was found to be safe in healthy and asthmatic subjects, and improvements in lung function were accompanied by increases in circulating anti-inflammatory markers in asthmatic subjects. Based on the results of this preliminary clinical study, we propose that this blend may improve lung function and inflammation by supplementing microbes in the gut to increase SCFA production in systemic circulation. Clinical relevance of these findings affects both patients with and without existing respiratory conditions who are seeking a means to support their lung health.

## Methods

### Trial Design

The study was conducted in accordance with the Declaration of Helsinki and approved by the Clinical Research Ethics Committee of the Cork Teaching Hospitals of University College Cork (ECM 4 (n) 7/9/2021& ECM 5 (7) 10/26/2021 & ECM 3 (lll) 11/16/2021).

This study was an open-label, exploratory pilot study to assess the safety of 4 weeks of dosing a probiotic and herbal blend (resB Lung Support, ResBiotic Nutrition) in healthy and asthmatic participants. Forty participants were screened to identify 22 eligible subjects. The study population consisted of n=11 healthy participants and n=11 participants diagnosed with asthma. Within these populations, active smoking status was noted (healthy smokers n=3, asthmatic smokers n=4). Informed consent was obtained from all subjects involved in the study. The study protocol consisted of four onsite visits over a 6-week period. Participants were pre-screened with an online questionnaire and invited for an onsite screening visit (Visit 1) to confirm their eligibility. A urine drug test was conducted and a urine test for pregnancy was performed for individuals of childbearing potential.

No changes were made to the methods after the trial started.

#### Participants

Male and female participants ages 18-65 were recruited who were either in general good health at the discretion of the investigator or had asthma and were on stable medication for at least 3 months. Exclusion criteria included pregnancy, acute or chronic illness which by the investigator’s judgment precluded them from participating in the study, ≤2 hospital admissions in the past 6 months, and use of antibiotics, probiotics, immunosuppressive medications, or oral steroids (>10 mg/day) for >3 days in the previous 12 weeks. Participants were also excluded if they had made any major dietary changes or changed medications or supplements in the 30 days prior to enrollment.

Participants were enrolled and conducted site visits at a single site: Atlantia Food Trials, Heron House Office, Blackpool Retail Park, in Cork, Ireland.

#### Interventions

All 22 participants took the supplement twice daily for 4 weeks (28 days). One capsule contains 8.25×10^9^ CFU *Lactobacillus plantarum*, 7.9×10^9^ CFU *Lactobacillus acidophilus*, 6.4×10^9^ CFU *Lactobacillus rhamnosus*, 48.0 mg vasaka (*Adhatoda vasica* root) extract, 42.0 mg holy basil (*Ocimum sanctum* leaf) extract, and 30.0 mg turmeric (*Curcuma longa* root) extract. At Visit 1, participants were provided with a 30-day supply plus two days of overage. Supplementation started after completion of Visit 2, where participants were instructed to consume one capsule twice daily with food and water at home. At Visits 3 and 4, participants returned all unused investigational product so that compliance could be calculated. Participants had to consume at least 75% (45 capsules) of their supply to be deemed compliant.

#### Outcomes

The primary objective of this trial was to measure safety of the blend in healthy and asthmatic study participants. Safety was measured by number of participants experiencing at least one adverse event (AE); number of AEs including causality, severity, and seriousness assessments; number of participants with discontinuations due to AEs; change in systolic blood pressure, diastolic blood pressure, heart rate, and body temperature from baseline to Week 2 and Week 4; change in blood safety parameters via serum chemistry profile and hematology profile from baseline to Week 2 and Week 4.

Exploratory objectives for this trial included: change in gut microbiota (16s) sequencing from baseline to Week 4; change in lung function measured by spirometry (Forced Expired Volume in 1 second (FEV1) and Forced Vital Capacity (FVC)) from baseline to Week 4; change in oxygen levels (% pulse oxygen levels) from baseline to Week 4; change in SGRQ score from baseline to Week 4.

At Baseline and Week 4, participants had blood drawn for biomarker analysis, spirometry measured, and completed the Saint George’s Respiratory Questionnaire (SGRQ). Participants supplied a stool sample at Visit 2 and Visit 4. Vitals, safety blood parameters, and adverse events/severe adverse events (AE/SAE) were monitored at each visit. Participants completed an end-of-study product questionnaire at Visit 4. There were no changes to the study protocol after the study initiated.

#### Blinding

This was an open label study where study personnel and the participants were aware of the product they were taking; blinding did not apply.

#### Statistical Methods

The primary method of analysis was descriptive statistics. Summary statistics for continuous measures were provided for the actual measurements at each visit and change from baseline to each visit. These tables included sample size, minimum and maximum statistics, mean, median, quartiles, and standard deviations. Key variables were categorized into clinical ranges. In the summary tables, counts and percentages were used in the frequency tables.

Inferential statistics were run to provide information on the trends within the data. Post-hoc paired t-tests were conducted to assess within group changes from baseline to Week 4 in the healthy and asthmatic populations separately. Participants were further segmented into smokers vs. non-smokers in assessing the changes from baseline to Week 4 in serum SCFA using paired t-tests. All analyses requiring significance testing were two-sided at a 5% significance level. Results were viewed as statistically significant if the p-value was less than or equal to 0.05.

### Microbiome Amplicon Sequencing and Analysis

Microbial DNA was extracted from human stool samples and 16S amplicon sequencing was performed using the Illumina MiSeq platform at the University of Alabama at Birmingham Microbiome Resource Core Facility under the direction of Dr. Casey Morrow. Sequencing data quality control, alignment and demultiplexing were perform by the using a custom script built using MOTHUR and QIIME 2 with amplicon sequence variants (ASVs) identified using SILVA. Processed ASV tables were imported into MicrobiomeAnalyst for further analysis and data visualization (38, 39). ASVs that appeared in less than two samples, were prevalent in 10% of samples and less than 5% of the inter quartile range were removed, leading to the removal of 504 low abundance features. Cumulative sum scaling was performed but not rarefication or transformation. Alpha diversity was quantified with the Shannon and Chao1 Indices. We visualized beta diversity with principal coordinates analysis of Bray-Curtis dissimilarity matrices and performed significance testing using permutational multivariate analysis of variance (PERMANOVA) and permutational multivariate analysis of dispersion (PERMDISP). Feature selection was performed using MetagenomeSeq and DESeq2.

### Qualitative PCR

DNA was re-extracted from same fecal material used in MiSeq with Zymobiomics DNA Mini-Prep kit (Zymo Research). PCR (Qiagen Fast Cycling PCR kit) was performed on samples with at least 3ng/uL total DNA by pico green quantification (Quant-It dsDNA Assay Kit, ThermoFisher). A common 16S gene-based forward primer was utilized with strain-specific reverse primers at a similar site (0.4uM), in a 25uL reaction with 30ng DNA template, using SYBR Green dye (LTi) to monitor in real time. Analysis of PCR reactions was performed on 2% agarose gels. Bands were gel extracted (Qiagen Qiaquick Gel Extaction Kit) and sequenced to confirm species identity. ZymoBiomics Gut Microbiome Standard (negative control) was used to demonstrate lack of nonspecific amplification.

### Biomarker Analysis

Serum SCFA levels were measured by GC-MS at Creative Proteomics (Shirley, NY). The following SCFAs were analyzed: acetic acid (C2:0), propionic acid (C3:0), butyric acid (C4:0), isobutyric acid (C4:0i), valeric acid (C5:0); isovaleric acid (C5:0i), hexanoic acid (C6:0). Free short chain fatty acids were derivatized using methyl chloroformate in 1-propanol yielding propyl esters before subsequent liquid-liquid extraction into hexane and analysis on a SLB-5ms (30×0.25×1.0μm) (Supelco) column and detection using GC-EI-MS in SIM-mode. Instrumental analysis was performed on an Agilent 7890 GC coupled to an Agilent 5977 MSD (Agilent Technologies). Quantification was performed against a 5-point calibration curve.

## Data Availability

The data that support the findings of this study are openly available in SRA at https://www.ncbi.nlm.nih.gov/bioproject/PRJNA922968, reference number PRJNA922968.

## Contributions

Study conception and strategy: CVL. Study design: NMW, LQ, TN, ZN, XX, AG, CVL. Manuscript preparation and interpretation: NMW, TN, KAW, NA, AG, CVL, BAH. Clinical trial design and execution: NMW, TD, AG, CVL. Clinical data analysis: TN, ZN, LQ, GD, CK, AG, CVL, BAH, KT, ME, IM. All authors reviewed, advised, and approved the final version submitted for publication and agree to be accountable for the work.

