## Supplemental Methods and Figures for "Clinical Trial of a Probiotic and Herbal Supplement for Lung and Gut Health"

**Supplemental Table 1.** Frequency table of participant Abnormal and Clinically Significant (ABCS) safety parameters at Baseline, Week 2, and Week 4 in the safety population.

|  | **Asthma**  **N** | **Healthy**  **N** |
| --- | --- | --- |
| **GGT**  Baseline  Week 2  Week 4 | -  -  1 | 1  1  1 |
| **GGT, Neutrophils**  Baseline  Week 2  Week 4 | 1  -  - | -  -  - |
| **Phosphate**  Baseline  Week 2  Week 4 | -  -  - | 1  1  - |
| **Vitamin D**  Baseline  Week 2  Week 4 | -  -  - | 1  1  - |
| **Monocytes**  Baseline  Week 2  Week 4 | -  1  1 | -  -  - |
| **Urea**  Baseline  Week 2  Week 4 | -  -  - | -  -  1 |

**Supplemental Table 2.** Summary descriptive statistics for vitals at Baseline, Week 2, and Week 4 in the safety population.

|  | **Health Status** | **N** | | **Mean** | **SEM** | **Mdn** | **SD** | **Min** | **Max** | **Percentiles** | | |
| --- | --- | --- | --- | --- | --- | --- | --- | --- | --- | --- | --- | --- |
|  |  | **Valid** | **Missing** |  |  |  |  |  |  | **25** | **50** | **75** |
| Systolic BP [mmHg]  Visit 2 | Healthy | 11 | 0 | 128.36 | 3.79 | 133.00 | 12.56 | 105.00 | 145.00 | 118.00 | 133.00 | 140.00 |
|  | Asthma | 11 | 0 | 126.18 | 3.54 | 125.00 | 11.74 | 109.00 | 152.00 | 117.00 | 125.00 | 130.00 |
| Diastolic BP [mmHg]  Visit 2 | Healthy | 11 | 0 | 75.00 | 2.29 | 76.00 | 7.59 | 62.00 | 86.00 | 70.00 | 76.00 | 81.00 |
|  | Asthma | 11 | 0 | 74.82 | 2.01 | 74.00 | 6.68 | 67.00 | 84.00 | 69.00 | 74.00 | 82.00 |
| Systolic BP [mmHg]  Visit 3 | Healthy | 10 | 1 | 129.60 | 4.19 | 129.50 | 13.26 | 106.00 | 150.00 | 120.25 | 129.50 | 140.50 |
|  | Asthma | 10 | 1 | 128.30 | 3.74 | 124.50 | 11.81 | 114.00 | 152.00 | 120.50 | 124.50 | 135.75 |
| Diastolic BP [mmHg]  Visit 3 | Healthy | 10 | 1 | 76.70 | 3.29 | 77.50 | 10.41 | 54.00 | 93.00 | 71.25 | 77.50 | 83.25 |
|  | Asthma | 10 | 1 | 76.80 | 2.22 | 76.00 | 7.02 | 68.00 | 90.00 | 70.50 | 76.00 | 81.75 |
| Systolic BP [mmHg]  Visit 4 | Healthy | 11 | 0 | 128.73 | 5.41 | 132.00 | 17.93 | 97.00 | 162.00 | 116.00 | 132.00 | 133.00 |
|  | Asthma | 11 | 0 | 122.82 | 5.59 | 118.00 | 18.54 | 101.00 | 160.00 | 105.00 | 118.00 | 136.00 |
| Diastolic BP [mmHg]  Visit 4 | Healthy | 11 | 0 | 77.55 | 2.92 | 76.00 | 9.70 | 61.00 | 94.00 | 70.00 | 76.00 | 83.00 |
|  | Asthma | 11 | 0 | 71.00 | 2.71 | 68.00 | 8.99 | 60.00 | 86.00 | 63.00 | 68.00 | 78.00 |
| ∆Change Systolic BP  Visit 3 | Healthy | 10 | 1 | 2.40 | 1.94 | 1.50 | 6.15 | -3.00 | 15.00 | -3.00 | 1.50 | 5.50 |
|  | Asthma | 10 | 1 | 1.20 | 2.73 | 1.00 | 8.64 | -12.00 | 14.00 | -6.25 | 1.00 | 7.75 |
| ∆Change Systolic BP  Visit 4 | Healthy | 11 | 0 | 0.36 | 4.32 | -4.00 | 14.33 | -23.00 | 26.00 | -8.00 | -4.00 | 11.00 |
|  | Asthma | 11 | 0 | -3.36 | 3.67 | -7.00 | 12.18 | -24.00 | 11.00 | -11.00 | -7.00 | 8.00 |
| ∆Change Diastolic BP  Visit 3 | Healthy | 10 | 1 | 2.80 | 2.41 | 3.00 | 7.63 | -16.00 | 11.00 | 1.25 | 3.00 | 7.75 |
|  | Asthma | 10 | 1 | 1.20 | 1.91 | 1.50 | 6.05 | -10.00 | 10.00 | -3.25 | 1.50 | 6.50 |
| ∆Change Diastolic BP  Visit 4 | Healthy | 11 | 0 | 2.55 | 2.94 | 1.00 | 9.75 | -15.00 | 20.00 | -4.00 | 1.00 | 10.00 |
|  | Asthma | 11 | 0 | -3.82 | 2.48 | -4.00 | 8.22 | -22.00 | 10.00 | -8.00 | -4.00 | 2.00 |
| Temperature [˚C]  Visit 2 | Healthy | 11 | 0 | 36.11 | 0.13 | 36.00 | 0.43 | 35.50 | 36.80 | 35.70 | 36.00 | 36.50 |
|  | Asthma | 11 | 0 | 36.05 | 0.08 | 36.10 | 0.28 | 35.60 | 36.60 | 36.00 | 36.10 | 36.20 |
| Temperature [˚C]  Visit 3 | Healthy | 10 | 1 | 35.95 | 0.18 | 35.70 | 0.57 | 35.50 | 37.20 | 35.58 | 35.70 | 36.17 |
|  | Asthma | 10 | 1 | 35.87 | 0.10 | 35.95 | 0.31 | 35.50 | 36.40 | 35.58 | 35.95 | 36.10 |
| Temperature [˚C]  Visit 4 | Healthy | 11 | 0 | 36.12 | 0.17 | 36.00 | 0.55 | 35.40 | 37.50 | 35.80 | 36.00 | 36.40 |
|  | Asthma | 11 | 0 | 36.23 | 0.13 | 36.20 | 0.44 | 35.60 | 36.90 | 35.80 | 36.20 | 36.60 |
| Pulse [Bpm]  Visit 2 | Healthy | 11 | 0 | 70.64 | 2.85 | 70.00 | 9.44 | 59.00 | 85.00 | 62.00 | 70.00 | 79.00 |
|  | Asthma | 11 | 0 | 72.45 | 2.75 | 78.00 | 9.11 | 55.00 | 80.00 | 65.00 | 78.00 | 80.00 |
| Pulse [Bpm]  Visit 3 | Healthy | 10 | 1 | 65.40 | 2.25 | 65.50 | 7.11 | 56.00 | 77.00 | 59.75 | 65.50 | 70.25 |
|  | Asthma | 10 | 1 | 73.30 | 4.34 | 68.50 | 13.73 | 57.00 | 101.00 | 65.75 | 68.50 | 81.75 |
| Pulse  [Bpm] Visit 4 | Healthy | 11 | 0 | 73.09 | 4.31 | 71.00 | 14.30 | 49.00 | 105.00 | 66.00 | 71.00 | 79.00 |
|  | Asthma | 11 | 0 | 73.55 | 2.10 | 74.00 | 6.98 | 61.00 | 84.00 | 67.00 | 74.00 | 78.00 |
| ∆Change Pulse  Visit 3 | Healthy | 10 | 1 | -4.60 | 1.55 | -6.00 | 4.90 | -10.00 | 5.00 | -8.50 | -6.00 | -0.50 |
|  | Asthma | 10 | 1 | 1.40 | 4.36 | -2.00 | 13.80 | -22.00 | 21.00 | -9.25 | -2.00 | 14.50 |
| ∆Change Pulse  Visit 4 | Healthy | 11 | 0 | 2.45 | 3.67 | 1.00 | 12.18 | -13.00 | 26.00 | -5.00 | 1.00 | 7.00 |
|  | Asthma | 11 | 0 | 1.09 | 1.97 | 4.00 | 6.52 | -13.00 | 8.00 | -5.00 | 4.00 | 6.00 |
| ∆Change Temp  Visit 3 | Healthy | 10 | 1 | -0.20 | 0.25 | -0.20 | 0.78 | -1.20 | 1.00 | -1.03 | -0.20 | 0.47 |
|  | Asthma | 10 | 1 | -0.22 | 0.12 | -0.15 | 0.37 | -0.70 | 0.40 | -0.55 | -0.15 | 0.03 |
| ∆Change Temp  Visit 4 | Healthy | 11 | 0 | 0.01 | 0.24 | 0.10 | 0.79 | -1.40 | 1.90 | -0.40 | 0.10 | 0.20 |
|  | Asthma | 11 | 0 | 0.18 | 0.11 | 0.10 | 0.36 | -0.30 | 0.80 | -0.20 | 0.10 | 0.50 |

**Supplemental Table 3.** Summary descriptive statistics for safety blood panel at Baseline and Week 4 per health status in the safety population.

| **Safety Blood Panel** | **Time** | **Health Status** | **N** | **Mean** | **SD** | **Min** | **Max** | **Median** |
| --- | --- | --- | --- | --- | --- | --- | --- | --- |
| **Sodium (mmol/L)** | Baseline | Asthma | 11 | 138 | 2.09 | 135 | 141 | 137 |
|  |  | Healthy | 11 | 138 | 3 | 132 | 142 | 139 |
|  | Week 4 | Asthma | 11 | 140 | 1.19 | 138 | 142 | 139 |
|  |  | Healthy | 11 | 139 | 2.77 | 131 | 142 | 139 |
| **Potassium (mmol/L)** | Baseline | Asthma | 11 | 4.31 | 0.401 | 3.9 | 5.1 | 4.2 |
|  |  | Healthy | 10 | 4.17 | 0.347 | 3.4 | 4.5 | 4.25 |
|  | Week 4 | Asthma | 10 | 4.4 | 0.485 | 3.7 | 5.2 | 4.35 |
|  |  | Healthy | 11 | 4.26 | 0.461 | 3.6 | 5.2 | 4.2 |
| **Chloride (mmol/L)** | Baseline | Asthma | 11 | 103 | 2.1 | 101 | 107 | 103 |
|  |  | Healthy | 11 | 103 | 2.34 | 99 | 109 | 103 |
|  | Week 4 | Asthma | 11 | 105 | 1.95 | 103 | 109 | 104 |
|  |  | Healthy | 11 | 105 | 2.8 | 99 | 110 | 104 |
| **Urea (mmol/L)** | Baseline | Asthma | 11 | 4.95 | 0.93 | 3.2 | 5.9 | 5.3 |
|  |  | Healthy | 11 | 4.69 | 1.42 | 2.4 | 7.6 | 4.6 |
|  | Week 4 | Asthma | 11 | 4.86 | 1.1 | 3.2 | 6.5 | 5 |
|  |  | Healthy | 11 | 5.33 | 1.57 | 3 | 8.8 | 5 |
| **Creatinine (umol/L)** | Baseline | Asthma | 11 | 76 | 13.5 | 62 | 97 | 69 |
|  |  | Healthy | 11 | 72 | 12.2 | 54 | 98 | 70 |
|  | Week 4 | Asthma | 11 | 81.4 | 16.6 | 62 | 120 | 78 |
|  |  | Healthy | 11 | 68.9 | 12.1 | 47 | 92 | 67 |
| **AST (IU/L)** | Baseline | Asthma | 11 | 19.5 | 3.75 | 14 | 25 | 19 |
|  |  | Healthy | 11 | 20.9 | 5.58 | 14 | 31 | 21 |
|  | Week 4 | Asthma | 11 | 20 | 6.37 | 14 | 36 | 21 |
|  |  | Healthy | 11 | 20.1 | 6.27 | 14 | 34 | 21 |
| **ALP (IU/L)** | Baseline | Asthma | 11 | 67.8 | 17.6 | 51 | 101 | 64 |
|  |  | Healthy | 11 | 75.7 | 20.7 | 54 | 114 | 67 |
|  | Week 4 | Asthma | 11 | 65.9 | 15.3 | 50 | 89 | 62 |
|  |  | Healthy | 11 | 71.6 | 22.4 | 44 | 110 | 64 |
| **GGT (IU/L)** | Baseline | Asthma | 11 | 18.5 | 11.9 | 8 | 50 | 14 |
|  |  | Healthy | 11 | 19.6 | 16.6 | 8 | 66 | 15 |
|  | Week 4 | Asthma | 11 | 21.1 | 12.8 | 10 | 54 | 18 |
|  |  | Healthy | 11 | 21.7 | 23.3 | 9 | 90 | 14 |
| **Total Protein (g/L)** | Baseline | Asthma | 11 | 69.11 | 3.11 | 66 | 76 | 68 |
|  |  | Healthy | 11 | 71.2 | 3.71 | 64 | 76 | 71 |
|  | Week 4 | Asthma | 11 | 70.5 | 3.62 | 65 | 76 | 71 |
|  |  | Healthy | 11 | 70.3 | 2.41 | 66 | 74 | 70 |
| **Total Bilirubin (umol/L)** | Baseline | Asthma | 11 | 11.2 | 3.42 | 5.9 | 17.5 | 10.8 |
|  |  | Healthy | 11 | 7.52 | 3.46 | 2.2 | 16 | 7.3 |
|  | Week 4 | Asthma | 11 | 12.3 | 4.11 | 5.6 | 20 | 11.7 |
|  |  | Healthy | 11 | 8.28 | 3.28 | 3 | 12.6 | 8.6 |
| **Albumin (g/L)** | Baseline | Asthma | 11 | 42.7 | 1.49 | 40 | 45 | 43 |
|  |  | Healthy | 11 | 43.6 | 1.5 | 41 | 46 | 44 |
|  | Week 4 | Asthma | 11 | 42.9 | 2.74 | 38 | 47 | 43 |
|  |  | Healthy | 11 | 42.9 | 1.3 | 41 | 45 | 43 |
| **Globulins (g/L)** | Baseline | Asthma | 11 | 26.4 | 2.42 | 24 | 32 | 26 |
|  |  | Healthy | 11 | 27.5 | 2.38 | 23 | 31 | 27 |
|  | Week 4 | Asthma | 11 | 27.5 | 2.21 | 25 | 33 | 27 |
|  |  | Healthy | 11 | 27.4 | 1.86 | 25 | 31 | 27 |
| **Calcium (mmol/L)** | Baseline | Asthma | 11 | 2.37 | 0.124 | 2.13 | 2.54 | 2.38 |
|  |  | Healthy | 11 | 2.33 | 0.113 | 2.18 | 2.49 | 2.3 |
|  | Week 4 | Asthma | 11 | 2.37 | 0.0987 | 2.21 | 2.53 | 2.38 |
|  |  | Healthy | 11 | 2.38 | 0.0534 | 2.31 | 2.48 | 2.37 |
| **Magnesium (mmol/L)** | Baseline | Asthma | 11 | 0.822 | 0.038 | 0.75 | 0.86 | 0.83 |
|  |  | Healthy | 11 | 0.815 | 0.066 | 0.72 | 0.92 | 0.81 |
|  | Week 4 | Asthma | 11 | 0.834 | 0.069 | 0.74 | 0.94 | 0.85 |
|  |  | Healthy | 11 | 0.788 | 0.045 | 0.73 | 0.87 | 0.78 |
| **Phosphate (mm/L)** | Baseline | Asthma | 11 | 1.12 | 0.178 | 0.73 | 1.42 | 1.13 |
|  |  | Healthy | 11 | 1.16 | 0.237 | 0.82 | 1.59 | 1.14 |
|  | Week 4 | Asthma | 11 | 1.15 | 0.174 | 0.87 | 1.4 | 1.16 |
|  |  | Healthy | 11 | 1.09 | 0.131 | 0.93 | 1.36 | 1.07 |
| **Uric Acid (umol/L** | Baseline | Asthma | 11 | 304 | 105 | 185 | 487 | 291 |
|  |  | Healthy | 11 | 260 | 61.9 | 137 | 337 | 280 |
|  | Week 4 | Asthma | 11 | 340 | 118 | 194 | 515 | 334 |
|  |  | Healthy | 11 | 285 | 50.7 | 214 | 380 | 286 |
| **Glucose (mmol/L)** | Baseline | Asthma | 11 | 4.65 | 0.808 | 3.1 | 5.9 | 4.8 |
|  |  | Healthy | 11 | 4.48 | 0.553 | 3.3 | 5.2 | 4.6 |
|  | Week 4 | Asthma | 11 | 4.13 | 0.863 | 2.8 | 5.4 | 4 |
|  |  | Healthy | 11 | 4.64 | 0.772 | 3.5 | 6.5 | 4.3 |
| **White Cell Count (10e9/L)** | Baseline | Asthma | 11 | 5.79 | 2 | 3.61 | 9.67 | 5.22 |
|  |  | Healthy | 11 | 6.38 | 2.06 | 3.84 | 10 | 5.51 |
|  | Week 4 | Asthma | 11 | 5.88 | 1.89 | 3.66 | 9.08 | 6.31 |
|  |  | Healthy | 11 | 5.95 | 1.57 | 3.75 | 8.73 | 5.71 |
| **Red Cell Count (10e12/L)** | Baseline | Asthma | 11 | 4.59 | 0.391 | 4.09 | 5.07 | 4.41 |
|  |  | Healthy | 11 | 4.66 | 0.351 | 4.19 | 5.26 | 4.61 |
|  | Week 4 | Asthma | 11 | 4.55 | 0.29 | 4.17 | 5.01 | 4.52 |
|  |  | Healthy | 11 | 4.53 | 0.287 | 4.22 | 5.08 | 4.45 |
| **Haemoglobin g/dL)** | Baseline | Asthma | 11 | 14.3 | 1.03 | 13.1 | 15.6 | 14.1 |
|  |  | Healthy | 11 | 14.2 | 1.04 | 12.7 | 15.6 | 13.8 |
|  | Week 4 | Asthma | 11 | 14.3 | 0.943 | 12.4 | 15.6 | 14.2 |
|  |  | Healthy | 11 | 13.8 | 0.851 | 12.2 | 14.9 | 14.1 |
| **Haematocrit (L/L)** | Baseline | Asthma | 11 | 0.433 | 0.031 | 0.391 | 0.471 | 0.426 |
|  |  | Healthy | 11 | 0.433 | 0.024 | 0.397 | 0.473 | 0.429 |
|  | Week 4 | Asthma | 11 | 0.428 | 0.025 | 0.376 | 0.47 | 0.428 |
|  |  | Healthy | 11 | 0.42 | 0.022 | 0.382 | 0.45 | 0.423 |
| **MCV (fL)** | Baseline | Asthma | 11 | 94.4 | 3.56 | 88.7 | 99.5 | 93.6 |
|  |  | Healthy | 11 | 93.1 | 4.06 | 86.1 | 100 | 92.5 |
|  | Week 4 | Asthma | 11 | 94.1 | 3.35 | 90.2 | 98.6 | 93.2 |
|  |  | Healthy | 11 | 92.9 | 3.59 | 88.6 | 99.5 | 92.8 |
| **MCH (pg)** | Baseline | Asthma | 11 | 31.2 | 1.1 | 29.7 | 32.9 | 31.5 |
|  |  | Healthy | 11 | 30.5 | 1.32 | 28.4 | 32.3 | 30.6 |
|  | Week 4 | Asthma | 11 | 31.4 | 1.3 | 29.7 | 33.5 | 31.7 |
|  |  | Healthy | 11 | 30.5 | 1.39 | 28.5 | 32.6 | 30.5 |
| **MCHC (g/dL)** | Baseline | Asthma | 11 | 33.1 | 0.488 | 32.2 | 33.8 | 33.2 |
|  |  | Healthy | 11 | 32.7 | 0.904 | 31.4 | 34.2 | 32.6 |
|  | Week 4 | Asthma | 11 | 33.4 | 0.656 | 32.5 | 34.8 | 33.5 |
|  |  | Healthy | 11 | 32.8 | 0.78 | 31.9 | 34.1 | 32.7 |
| **RDW (%)** | Baseline | Asthma | 11 | 12.3 | 0.428 | 11.6 | 13.1 | 12.2 |
|  |  | Healthy | 11 | 12.5 | 0.688 | 11.8 | 13.7 | 12.2 |
|  | Week 4 | Asthma | 11 | 12.4 | 0.437 | 11.6 | 13.1 | 12.4 |
|  |  | Healthy | 11 | 12.5 | 0.757 | 11.6 | 13.9 | 12.2 |
| **Platelets (10e9/L)** | Baseline | Asthma | 11 | 256 | 60.5 | 181 | 396 | 234 |
|  |  | Healthy | 11 | 258 | 63 | 174 | 381 | 238 |
|  | Week 4 | Asthma | 11 | 264 | 65.9 | 175 | 419 | 245 |
|  |  | Healthy | 11 | 234 | 52.8 | 159 | 337 | 221 |
| **Neutrophils (10e9/L)** | Baseline | Asthma | 11 | 3.22 | 1.63 | 1.96 | 7.05 | 2.61 |
|  |  | Healthy | 11 | 3.67 | 1.43 | 2 | 6.57 | 2.91 |
|  | Week 4 | Asthma | 11 | 3.33 | 1.59 | 1.6 | 6.34 | 2.61 |
|  |  | Healthy | 11 | 3.36 | 1.36 | 1.84 | 6.45 | 3.13 |
| **Lymphocytes (10e9/L)** | Baseline | Asthma | 11 | 1.75 | 0.422 | 1.09 | 2.52 | 1.83 |
|  |  | Healthy | 11 | 2.04 | 0.677 | 1.17 | 3.74 | 2.02 |
|  | Week 4 | Asthma | 11 | 1.75 | 0.477 | 1.19 | 2.64 | 1.62 |
|  |  | Healthy | 11 | 1.86 | 0.521 | 0.92 | 2.51 | 1.98 |
| **Monocytes (10e9/L)** | Baseline | Asthma | 11 | 0.517 | 0.254 | 0.24 | 1.19 | 0.48 |
|  |  | Healthy | 11 | 0.436 | 0.128 | 0.32 | 0.66 | 0.42 |
|  | Week 4 | Asthma | 11 | 0.524 | 0.204 | 0.24 | 0.95 | 0.56 |
|  |  | Healthy | 11 | 0.466 | 0.187 | 0.3 | 0.99 | 0.42 |
| **Eosinophils (10e9/L)** | Baseline | Asthma | 11 | 0.254 | 0.241 | 0.04 | 0.92 | 0.2 |
|  |  | Healthy | 11 | 0.188 | 0.093 | 0.06 | 0.38 | 0.2 |
|  | Week 4 | Asthma | 11 | 0.241 | 0.17 | 0.04 | 0.62 | 0.19 |
|  |  | Healthy | 11 | 0.214 | 0.123 | 0.06 | 0.43 | 0.16 |
| **Basophils (10e9/L)** | Baseline | Asthma | 11 | 0.047 | 0.016 | 0.03 | 0.07 | 0.05 |
|  |  | Healthy | 11 | 0.046 | 0.022 | 0.02 | 0.08 | 0.04 |
|  | Week 4 | Asthma | 11 | 0.047 | 0.014 | 0.02 | 0.07 | 0.05 |
|  |  | Healthy | 11 | 0.045 | 0.027 | 0.01 | 0.1 | 0.04 |

**Supplementary Table 4.** Deseq2 Analysis of Probiotic Exposure.

|  | **log2FC** | **lfcSE** | **Pvalues** | **FDR** |
| --- | --- | --- | --- | --- |
| **uncultured_bacterium_58** | 24.961 | 2.2212 | 2.667E-29 | 9.4349E-27 |
| **human_gut_metagenome_3** | 24.791 | 2.2159 | 4.6823E-29 | 9.4349E-27 |
| **uncultured_organism_49** | 24.206 | 2.3862 | 3.5216E-24 | 4.7307E-22 |
| **D\_5\_\_Prevotella_9_unclassified** | -26.553 | 2.9181 | 9.084E-20 | 9.1521E-18 |
| **uncultured_organism_20** | 25.538 | 2.9181 | 2.1022E-18 | 1.6106E-16 |
| **uncultured_organism_13** | 25.495 | 2.9181 | 2.3979E-18 | 1.6106E-16 |
| **uncultured_bacterium_13** | -24.997 | 2.9183 | 1.0737E-17 | 6.1813E-16 |
| **uncultured_rumen_bacterium_2** | -23.984 | 2.9187 | 2.082E-16 | 1.0488E-14 |
| **unidentified_7** | -23.673 | 2.9188 | 5.0462E-16 | 2.2596E-14 |
| **uncultured_organism_59** | 23.299 | 2.9187 | 1.4296E-15 | 5.7612E-14 |
| **uncultured_bacterium_69** | 22.852 | 2.9185 | 4.8711E-15 | 1.7846E-13 |
| **uncultured_organism_38** | -22.685 | 2.9187 | 7.694E-15 | 2.5839E-13 |
| **uncultured_bacterium_89** | 22.563 | 2.9191 | 1.082E-14 | 3.3541E-13 |
| **D\_5\_\_Tyzzerella_4_unclassified** | 22.306 | 2.9 | 1.4518E-14 | 4.1791E-13 |
| **metagenome_9** | 21.992 | 2.9197 | 4.9835E-14 | 1.3389E-12 |
| **uncultured_bacterium_35** | 6.4548 | 1.8067 | 0.00035342 | 0.0089017 |
| **D\_5\_\_Blautia_unclassified** | -1.4247 | 0.40958 | 0.00050446 | 0.011959 |
| **uncultured_organism_2** | -2.1891 | 0.64977 | 0.00075428 | 0.016888 |
| **unidentified_10** | -5.6439 | 1.688 | 0.00082729 | 0.017547 |
| **Bifidobacterium_bifidum** | 5.2634 | 1.5953 | 0.00096928 | 0.019531 |
| **unidentified_4** | -7.8851 | 2.4888 | 0.0015336 | 0.02943 |
| **uncultured_bacterium_173** | 5.7613 | 1.8518 | 0.0018637 | 0.034139 |
| **uncultured_Ruminococcaceae_bacterium_1** | 4.8992 | 1.6578 | 0.003124 | 0.054738 |
| **unidentified_6** | -5.3165 | 1.8211 | 0.0035072 | 0.058892 |
| **uncultured_spirochete** | 4.4497 | 1.5331 | 0.0037032 | 0.059695 |
| **Clostridium_sp__K4410_MGS_306** | -4.4277 | 1.5946 | 0.0054924 | 0.085132 |
| **human_gut_metagenome_4** | 4.3651 | 1.6373 | 0.0076729 | 0.11453 |
| **metagenome_15** | 4.1641 | 1.5899 | 0.0088169 | 0.12365 |
| **uncultured_Bacteroides_sp_** | -7.6346 | 2.9185 | 0.0088975 | 0.12365 |
| **unidentified_3** | 3.1736 | 1.2654 | 0.012143 | 0.16312 |
| **gut_metagenome_46** | 5.5642 | 2.2829 | 0.014798 | 0.19237 |
| **D\_5\_\_Ruminiclostridium_6_unclassified** | -3.9973 | 1.6483 | 0.015305 | 0.19274 |
| **D\_5\_\_Christensenellaceae_R_7_group_unclassified** | 3.1429 | 1.3421 | 0.01919 | 0.23152 |
| **D\_4\_\_Lachnospiraceae_unclassified_12** | -4.7265 | 2.03 | 0.019893 | 0.23152 |
| **D\_4\_\_Lachnospiraceae_unclassified_4** | -4.665 | 2.007 | 0.020107 | 0.23152 |
| **uncultured_bacterium_193** | -5.8246 | 2.5545 | 0.022599 | 0.25298 |
| **Bifidobacterium_longum_subsp__longum_1** | 2.1645 | 0.95655 | 0.023647 | 0.25756 |
| **gut_metagenome_25** | 5.7746 | 2.5702 | 0.024656 | 0.2582 |
| **Bacteroides_ovatus_V975_2** | -3.8641 | 1.7282 | 0.025355 | 0.2582 |
| **D\_5\_\_uncultured_unclassified_5** | -4.053 | 1.816 | 0.025628 | 0.2582 |
| **uncultured_organism_5** | -1.2792 | 0.58001 | 0.027425 | 0.26527 |
| **uncultured_bacterium_49** | 3.7023 | 1.6811 | 0.027646 | 0.26527 |
| **uncultured_organism_1** | -0.76201 | 0.35002 | 0.029479 | 0.27154 |
| **uncultured_bacterium_109** | 3.7994 | 1.7522 | 0.030133 | 0.27154 |
| **D\_5\_\_Streptococcus_unclassified** | -4.8706 | 2.2488 | 0.03032 | 0.27154 |
| **D\_4\_\_Ruminococcaceae_unclassified_6** | 4.7624 | 2.2367 | 0.033239 | 0.28805 |
| **uncultured_bacterium_70** | -6.2031 | 2.9192 | 0.033594 | 0.28805 |
| **uncultured_bacterium_147** | -5.9031 | 2.9195 | 0.043183 | 0.36256 |
| **metagenome_25** | -4.4938 | 2.2497 | 0.045771 | 0.37073 |
| **gut_metagenome_58** | 5.0863 | 2.5538 | 0.046404 | 0.37073 |
| **bacterium_YE57** | 3.3444 | 1.6831 | 0.046916 | 0.37073 |
| **uncultured_organism_36** | -3.5018 | 1.7772 | 0.048786 | 0.37809 |
| **uncultured_bacterium_131** | 5.6337 | 2.92 | 0.053691 | 0.4057 |
| **uncultured_bacterium_164** | 5.5848 | 2.9028 | 0.054362 | 0.4057 |
| **uncultured_bacterium_82** | 5.5646 | 2.9201 | 0.056702 | 0.41547 |
| **uncultured_rumen_bacterium_3** | 4.1033 | 2.166 | 0.058166 | 0.41858 |
| **uncultured_bacterium_61** | -2.4023 | 1.2769 | 0.059911 | 0.41946 |
| **uncultured_bacterium_159** | 3.158 | 1.6896 | 0.061606 | 0.41946 |
| **uncultured_bacterium_40** | -1.4002 | 0.75116 | 0.062307 | 0.41946 |
| **uncultured_bacterium_5** | -0.7988 | 0.42875 | 0.062451 | 0.41946 |
| **uncultured_bacterium_98** | -4.4114 | 2.3825 | 0.064087 | 0.41958 |
| **uncultured_organism_25** | 2.6995 | 1.4635 | 0.065105 | 0.41958 |
| **gut_metagenome_19** | -5.3767 | 2.9202 | 0.065592 | 0.41958 |
| **uncultured_organism_53** | -2.3002 | 1.2584 | 0.067579 | 0.42553 |
| **gut_metagenome_3** | 1.7494 | 0.96551 | 0.070002 | 0.43401 |
| **uncultured_organism** | -0.58966 | 0.32755 | 0.071823 | 0.43855 |
| **uncultured_organism_35** | 5.2333 | 2.9207 | 0.073161 | 0.44006 |
| **uncultured_bacterium_180** | 2.216 | 1.2512 | 0.076556 | 0.45 |
| **uncultured_bacterium_235** | -4.545 | 2.5711 | 0.077103 | 0.45 |
| **gut_metagenome_20** | 5.1449 | 2.9208 | 0.078164 | 0.45 |
| **uncultured_bacterium_66** | -1.2906 | 0.74168 | 0.081833 | 0.45675 |
| **uncultured_organism_63** | 5.0447 | 2.9211 | 0.084163 | 0.45675 |
| **gut_metagenome_17** | 5.0406 | 2.9211 | 0.084417 | 0.45675 |
| **uncultured_bacterium_87** | 2.469 | 1.443 | 0.087067 | 0.45675 |
| **uncultured_bacterium_170** | -4.9801 | 2.9209 | 0.088194 | 0.45675 |
| **uncultured_bacterium_138** | 3.641 | 2.1416 | 0.089109 | 0.45675 |
| **uncultured_bacterium_33** | -4.9577 | 2.9209 | 0.089644 | 0.45675 |
| **uncultured_organism_84** | 4.3695 | 2.5763 | 0.08988 | 0.45675 |
| **uncultured_organism_95** | -4.4716 | 2.6391 | 0.090189 | 0.45675 |
| **uncultured_rumen_bacterium_4** | 3.1295 | 1.8497 | 0.09067 | 0.45675 |
| **uncultured_organism_23** | 1.4267 | 0.84858 | 0.092705 | 0.45758 |
| **gut_metagenome_11** | 1.672 | 1.0013 | 0.094936 | 0.45758 |
| **gut_metagenome_28** | 4.5118 | 2.7034 | 0.095123 | 0.45758 |
| **uncultured_organism_26** | -0.37666 | 0.22603 | 0.095629 | 0.45758 |
| **uncultured_bacterium_84** | 4.8478 | 2.9215 | 0.097045 | 0.45758 |
| **uncultured_bacterium_94** | 3.5137 | 2.1213 | 0.097648 | 0.45758 |
| **human_gut_metagenome** | 3.1437 | 1.9224 | 0.10199 | 0.46072 |
| **uncultured_bacterium_127** | -3.1355 | 1.926 | 0.10353 | 0.46072 |
| **uncultured_bacterium_80** | -4.7547 | 2.9214 | 0.10362 | 0.46072 |
| **uncultured_bacterium_198** | 4.7489 | 2.9218 | 0.10409 | 0.46072 |
| **uncultured_bacterium_25** | -3.1016 | 1.9097 | 0.10435 | 0.46072 |
| **uncultured_bacterium_37** | 4.5903 | 2.8334 | 0.10522 | 0.46072 |
| **gut_metagenome_22** | 2.4864 | 1.5396 | 0.10632 | 0.46072 |
| **uncultured_organism_29** | -1.085 | 0.6766 | 0.10881 | 0.46608 |
| **D\_5\_\_Ruminococcaceae_UCG_010_unclassified_1** | 4.6716 | 2.922 | 0.10987 | 0.46608 |
| **uncultured_organism_69** | -4.6451 | 2.9217 | 0.11186 | 0.46959 |
| **Streptococcus_salivarius_subsp__thermophilus** | -1.8251 | 1.1607 | 0.11587 | 0.48138 |
| **uncultured_organism_8** | -2.124 | 1.3619 | 0.11887 | 0.48883 |
| **uncultured_organism_61** | 1.9156 | 1.2341 | 0.12061 | 0.49096 |
| **uncultured_organism_27** | -1.6836 | 1.1023 | 0.12667 | 0.5077 |
| **uncultured_bacterium_73** | 4.4572 | 2.9226 | 0.12724 | 0.5077 |
| **gut_metagenome_49** | -2.3421 | 1.5412 | 0.12861 | 0.50814 |
| **uncultured_organism_7** | -1.8509 | 1.2249 | 0.13077 | 0.51166 |
| **D\_4\_\_Lachnospiraceae_unclassified_2** | -2.8127 | 1.8907 | 0.13685 | 0.52116 |
| **gut_metagenome_10** | 2.0588 | 1.3841 | 0.13689 | 0.52116 |
| **uncultured_bacterium_156** | 4.3458 | 2.923 | 0.13708 | 0.52116 |
| **uncultured_Firmicutes_bacterium** | -1.9047 | 1.2899 | 0.13977 | 0.52273 |
| **Slackia_sp__NATTS** | 3.0837 | 2.0967 | 0.14135 | 0.52273 |
| **uncultured_organism_104** | 3.8169 | 2.6066 | 0.14311 | 0.52273 |
| **Parabacteroides_johnsonii_CL02T12C29** | 4.2755 | 2.9232 | 0.14357 | 0.52273 |
| **bacterium_YE57_1** | 4.2525 | 2.9233 | 0.14575 | 0.52273 |
| **uncultured_bacterium_142** | 4.2461 | 2.9233 | 0.14637 | 0.52273 |
| **uncultured_Eubacterium_sp_** | -0.84669 | 0.5845 | 0.14745 | 0.52273 |
| **D\_5\_\_Alistipes_unclassified_2** | 1.7995 | 1.2435 | 0.14787 | 0.52273 |
| **D\_5\_\_Alistipes_unclassified** | -1.1541 | 0.80131 | 0.1498 | 0.52495 |
| **uncultured_rumen_bacterium** | 1.4641 | 1.0326 | 0.15622 | 0.53131 |
| **uncultured_Alistipes_sp_** | -0.7513 | 0.53181 | 0.15774 | 0.53131 |
| **gut_metagenome_44** | -4.1287 | 2.9233 | 0.15784 | 0.53131 |
| **uncultured_bacterium_132** | 2.8783 | 2.0486 | 0.16002 | 0.53131 |
| **metagenome_14** | 4.1077 | 2.9239 | 0.16006 | 0.53131 |
| **D\_5\_\_Blautia_unclassified_3** | -4.1008 | 2.9234 | 0.16069 | 0.53131 |
| **uncultured_bacterium_39** | 0.7369 | 0.52551 | 0.16084 | 0.53131 |
| **D\_5\_\_uncultured_unclassified_1** | -3.0054 | 2.1597 | 0.16406 | 0.53752 |
| **D\_5\_\_Alistipes_unclassified_3** | 3.4297 | 2.4992 | 0.16997 | 0.54671 |
| **human_gut_metagenome_12** | -4.0055 | 2.9237 | 0.17068 | 0.54671 |
| **D\_4\_\_Ruminococcaceae_unclassified_10** | 4.004 | 2.9243 | 0.17093 | 0.54671 |
| **Clostridium_sp__CAG:306** | -3.5835 | 2.6449 | 0.17545 | 0.55232 |
| **Bacteroides_dorei_1** | -1.4922 | 1.1042 | 0.17656 | 0.55232 |
| **uncultured_organism_9** | 0.9543 | 0.70653 | 0.1768 | 0.55232 |
| **uncultured_organism_40** | -1.6865 | 1.2616 | 0.18129 | 0.56001 |
| **D\_3\_\_Clostridiales_unclassified_1** | 2.1453 | 1.6076 | 0.18204 | 0.56001 |
| **gut_metagenome_24** | 1.2807 | 0.96912 | 0.18632 | 0.56241 |
| **gut_metagenome_60** | 3.8618 | 2.925 | 0.18674 | 0.56241 |
| **D\_4\_\_Clostridiales_vadinBB60_group_unclassified_2** | 3.8586 | 2.925 | 0.1871 | 0.56241 |
| **uncultured_bacterium_187** | 3.4733 | 2.6406 | 0.1884 | 0.56241 |
| **uncultured_bacterium_269** | 3.4426 | 2.6332 | 0.19109 | 0.56623 |
| **uncultured_bacterium_121** | 2.2044 | 1.7086 | 0.19697 | 0.57765 |
| **uncultured_organism_58** | 1.782 | 1.3837 | 0.19781 | 0.57765 |
| **uncultured_organism_81** | -3.7034 | 2.9251 | 0.20548 | 0.59573 |
| **uncultured_bacterium_81** | -1.3357 | 1.0603 | 0.20776 | 0.59806 |
| **uncultured_bacterium_210** | 3.6623 | 2.926 | 0.21069 | 0.59857 |
| **uncultured_bacterium_115** | 2.2573 | 1.8043 | 0.21091 | 0.59857 |
| **uncultured_organism_41** | -1.2337 | 0.99092 | 0.21314 | 0.59888 |
| **uncultured_bacterium_12** | -3.1197 | 2.5175 | 0.21527 | 0.59888 |
| **Bacteroides_fragilis** | 2.2634 | 1.8273 | 0.21548 | 0.59888 |
| **Ruminococcus_sp__DJF_VR70k1** | 2.3019 | 1.877 | 0.22006 | 0.60744 |
| **metagenome_5** | 1.6802 | 1.386 | 0.2254 | 0.61601 |
| **D\_5\_\_GCA_900066225_unclassified** | 3.5303 | 2.9267 | 0.22773 | 0.61601 |
| **unidentified_14** | 3.5286 | 2.9268 | 0.22796 | 0.61601 |
| **Streptococcus_pneumoniae** | 3.5186 | 2.9268 | 0.22929 | 0.61601 |
| **uncultured_organism_46** | 2.0586 | 1.7417 | 0.23723 | 0.62563 |
| **D\_4\_\_Lachnospiraceae_unclassified_13** | -3.4574 | 2.9264 | 0.23742 | 0.62563 |
| **uncultured_Clostridium_sp__2** | 1.8586 | 1.5762 | 0.23834 | 0.62563 |
| **uncultured_bacterium_154** | 3.2159 | 2.7316 | 0.23908 | 0.62563 |
| **uncultured_bacterium_101** | -2.0271 | 1.7355 | 0.2428 | 0.6283 |
| **unidentified_9** | 3.4017 | 2.9275 | 0.24525 | 0.6283 |
| **uncultured_bacterium_149** | 1.6498 | 1.4211 | 0.24567 | 0.6283 |
| **uncultured_organism_17** | 0.85226 | 0.73515 | 0.24633 | 0.6283 |
| **uncultured_bacterium_197** | 1.9585 | 1.6991 | 0.24903 | 0.63119 |
| **uncultured_bacterium_68** | 3.2805 | 2.8982 | 0.25767 | 0.64767 |
| **Anaerostipes_hadrus** | 0.82171 | 0.72759 | 0.25875 | 0.64767 |
| **uncultured_bacterium_75** | -2.0828 | 1.8659 | 0.2643 | 0.65749 |
| **uncultured_Roseburia_sp_** | 1.9452 | 1.7563 | 0.26804 | 0.65902 |
| **metagenome_4** | -0.95021 | 0.85941 | 0.26888 | 0.65902 |
| **D\_5\_\_Oxalobacter_unclassified** | 1.5198 | 1.3773 | 0.26982 | 0.65902 |
| **uncultured_organism_11** | -0.41127 | 0.3747 | 0.27238 | 0.66057 |
| **Bacteroides_thetaiotaomicron** | -0.5519 | 0.50425 | 0.27374 | 0.66057 |
| **D\_4\_\_Lachnospiraceae_unclassified_14** | -2.9339 | 2.6917 | 0.27574 | 0.66144 |
| **uncultured_organism_15** | -1.3725 | 1.2806 | 0.2838 | 0.67675 |
| **uncultured_bacterium_67** | 1.6546 | 1.5674 | 0.29113 | 0.69014 |
| **uncultured_bacterium_8** | 1.1798 | 1.1418 | 0.30147 | 0.71049 |
| **uncultured_bacterium_178** | -3.0088 | 2.9294 | 0.30438 | 0.71256 |
| **uncultured_organism_88** | 2.9999 | 2.9299 | 0.30589 | 0.71256 |
| **uncultured_bacterium_172** | -2.8077 | 2.7857 | 0.3135 | 0.72312 |
| **uncultured_bacterium_215** | 1.6941 | 1.6826 | 0.31401 | 0.72312 |
| **D\_2\_\_Clostridia_unclassified** | 2.3915 | 2.4129 | 0.32162 | 0.73643 |
| **uncultured_bacterium_15** | 1.8506 | 1.9036 | 0.33099 | 0.74669 |
| **uncultured_bacterium_1** | -0.29924 | 0.30812 | 0.33146 | 0.74669 |
| **uncultured_organism_48** | -1.1349 | 1.1708 | 0.33239 | 0.74669 |
| **uncultured_organism_34** | 1.6345 | 1.6901 | 0.33351 | 0.74669 |
| **metagenome_1** | 2.7585 | 2.8862 | 0.33919 | 0.75295 |
| **D\_4\_\_Lachnospiraceae_unclassified_3** | 1.3961 | 1.464 | 0.34027 | 0.75295 |
| **gut_metagenome_52** | -1.5804 | 1.6629 | 0.34191 | 0.75295 |
| **uncultured_bacterium_28** | 0.93013 | 1.001 | 0.35278 | 0.76997 |
| **gut_metagenome_7** | 1.6248 | 1.7511 | 0.35346 | 0.76997 |
| **uncultured_organism_12** | 1.1145 | 1.2084 | 0.35636 | 0.77067 |
| **uncultured_bacterium_55** | 1.1944 | 1.2984 | 0.3576 | 0.77067 |
| **metagenome_18** | 2.6485 | 2.9212 | 0.36459 | 0.78154 |
| **uncultured_bacterium_42** | -1.5884 | 1.7625 | 0.36747 | 0.78335 |
| **human_gut_metagenome_2** | -1.8299 | 2.0384 | 0.36932 | 0.78335 |
| **uncultured_Clostridium_sp__1** | 1.8276 | 2.0631 | 0.37569 | 0.79269 |
| **uncultured_bacterium_249** | 2.5572 | 2.9332 | 0.3833 | 0.80442 |
| **metagenome_13** | 1.3924 | 1.6061 | 0.38599 | 0.80442 |
| **uncultured_bacterium_14** | 0.22712 | 0.26267 | 0.38724 | 0.80442 |
| **uncultured_bacterium_21** | -0.93963 | 1.0971 | 0.39176 | 0.80964 |
| **uncultured_archaeon** | 1.1928 | 1.4007 | 0.39446 | 0.81058 |
| **uncultured_rumen_bacterium_1** | 1.6586 | 1.955 | 0.39624 | 0.81058 |
| **uncultured_bacterium_107** | -0.99273 | 1.1829 | 0.40134 | 0.81687 |
| **uncultured_rumen_bacterium_5** | 1.0553 | 1.2796 | 0.40955 | 0.8238 |
| **uncultured_bacterium_27** | 0.49102 | 0.60488 | 0.41693 | 0.8238 |
| **uncultured_Ruminococcus_sp_** | -0.84162 | 1.0375 | 0.41723 | 0.8238 |
| **gut_metagenome_35** | 1.9948 | 2.4603 | 0.41748 | 0.8238 |
| **Faecalibacterium_prausnitzii** | 1.5476 | 1.9089 | 0.41752 | 0.8238 |
| **gut_metagenome_9** | -0.5812 | 0.72888 | 0.42523 | 0.8238 |
| **Ruminococcus_sp__UNK_MGS_30** | -1.8314 | 2.298 | 0.42547 | 0.8238 |
| **Bacteroides_ovatus_V975** | -1.0689 | 1.3415 | 0.42556 | 0.8238 |
| **uncultured_organism_10** | -0.27809 | 0.34987 | 0.42671 | 0.8238 |
| **bacterium_enrichment_culture_clone_LA92** | -0.83102 | 1.0568 | 0.43167 | 0.8238 |
| **uncultured_bacterium_134** | 2.2761 | 2.9113 | 0.43432 | 0.8238 |
| **uncultured_bacterium_4** | -0.54371 | 0.69626 | 0.43486 | 0.8238 |
| **uncultured_organism_64** | 1.7734 | 2.2785 | 0.43637 | 0.8238 |
| **Lactobacillus_gasseri** | 0.46534 | 0.59956 | 0.43767 | 0.8238 |
| **gut_metagenome_26** | -1.5112 | 1.9517 | 0.43876 | 0.8238 |
| **uncultured_Blautia_sp_** | -0.24241 | 0.31367 | 0.43962 | 0.8238 |
| **uncultured_bacterium_16** | -0.37423 | 0.48586 | 0.44115 | 0.8238 |
| **uncultured_bacterium_43** | 1.5039 | 1.9608 | 0.44311 | 0.8238 |
| **uncultured_organism_50** | 2.217 | 2.8937 | 0.44359 | 0.8238 |
| **uncultured_bacterium_125** | -1.1942 | 1.5731 | 0.44778 | 0.82777 |
| **uncultured_organism_21** | -1.1831 | 1.5839 | 0.45509 | 0.83745 |
| **metagenome_8** | -1.2657 | 1.7131 | 0.45999 | 0.8383 |
| **D\_3\_\_Clostridiales_unclassified_5** | 1.6042 | 2.1791 | 0.46161 | 0.8383 |
| **uncultured_organism_22** | 2.1243 | 2.8867 | 0.46179 | 0.8383 |
| **Bacteroides_ovatus_V975_3** | 1.3652 | 1.8647 | 0.4641 | 0.8387 |
| **uncultured_organism_32** | -1.0278 | 1.4144 | 0.46743 | 0.83909 |
| **human_gut_metagenome_6** | -1.0468 | 1.4441 | 0.46853 | 0.83909 |
| **unidentified_2** | -0.70061 | 0.97094 | 0.47055 | 0.83909 |
| **uncultured_bacterium_238** | 2.0989 | 2.9249 | 0.47301 | 0.83975 |
| **D\_5\_\_Ruminiclostridium_9_unclassified** | -1.1095 | 1.5683 | 0.47928 | 0.84623 |
| **uncultured_Veillonellaceae_bacterium** | 1.4526 | 2.0607 | 0.48086 | 0.84623 |
| **D\_5\_\_Parabacteroides_unclassified** | -0.47719 | 0.68245 | 0.48441 | 0.84876 |
| **uncultured_bacterium_3** | 0.42105 | 0.60667 | 0.48766 | 0.85077 |
| **metagenome_7** | -0.93945 | 1.3709 | 0.49317 | 0.85197 |
| **uncultured_organism_45** | -0.52033 | 0.76089 | 0.49407 | 0.85197 |
| **Dorea_formicigenerans_ATCC_27755** | -0.21485 | 0.31463 | 0.49469 | 0.85197 |
| **uncultured_organism_14** | 0.45363 | 0.67978 | 0.50457 | 0.86352 |
| **uncultured_bacterium_51** | 0.78669 | 1.1825 | 0.50587 | 0.86352 |
| **uncultured_bacterium_257** | 1.9405 | 2.9302 | 0.50783 | 0.86352 |
| **gut_metagenome_2** | -0.44654 | 0.68574 | 0.51493 | 0.87192 |
| **uncultured_Thermoanaerobacterales_bacterium_1** | 1.8012 | 2.7991 | 0.51991 | 0.87666 |
| **uncultured_bacterium_232** | 1.086 | 1.7243 | 0.52883 | 0.88645 |
| **uncultured_bacterium_110** | -1.8064 | 2.8961 | 0.5328 | 0.88645 |
| **uncultured_bacterium_144** | 0.84586 | 1.359 | 0.53365 | 0.88645 |
| **Bacteroides_dorei** | -0.90512 | 1.4644 | 0.53653 | 0.88645 |
| **uncultured_bacterium_54** | -0.71438 | 1.1563 | 0.53671 | 0.88645 |
| **uncultured_bacterium_90** | -0.57182 | 0.93509 | 0.54086 | 0.88817 |
| **uncultured_bacterium_79** | -0.95398 | 1.5651 | 0.54216 | 0.88817 |
| **D\_5\_\_Methylobacterium_unclassified** | 1.7758 | 2.9414 | 0.54602 | 0.89088 |
| **uncultured_bacterium_31** | -1.3468 | 2.2585 | 0.55096 | 0.89133 |
| **uncultured_bacterium_32** | 0.70285 | 1.1831 | 0.55247 | 0.89133 |
| **uncultured_bacterium_45** | 0.31684 | 0.53397 | 0.55294 | 0.89133 |
| **uncultured_organism_31** | -1.6894 | 2.9001 | 0.5602 | 0.89872 |
| **uncultured_bacterium_200** | -1.437 | 2.4887 | 0.56367 | 0.89872 |
| **uncultured_bacterium_24** | -0.2318 | 0.40201 | 0.56421 | 0.89872 |
| **uncultured_bacterium_166** | 0.99211 | 1.7853 | 0.57842 | 0.90831 |
| **unidentified_1** | -0.45103 | 0.81488 | 0.57993 | 0.90831 |
| **uncultured_organism_47** | 1.2697 | 2.3071 | 0.58207 | 0.90831 |
| **Alistipes_indistinctus_YIT_12060** | -0.82408 | 1.5004 | 0.58285 | 0.90831 |
| **uncultured_organism_30** | 0.57825 | 1.0537 | 0.58316 | 0.90831 |
| **uncultured_Clostridiales_bacterium** | 1.0043 | 1.8329 | 0.58376 | 0.90831 |
| **uncultured_bacterium_85** | -0.79406 | 1.4783 | 0.59117 | 0.91369 |
| **uncultured_organism_55** | 1.1173 | 2.105 | 0.59557 | 0.91369 |
| **uncultured_bacterium_184** | -1.3646 | 2.571 | 0.59559 | 0.91369 |
| **uncultured_organism_4** | 0.54447 | 1.0406 | 0.60082 | 0.91369 |
| **metagenome_11** | -0.56437 | 1.0954 | 0.6064 | 0.91369 |
| **uncultured_bacterium_2** | -0.24801 | 0.48471 | 0.60888 | 0.91369 |
| **uncultured_bacterium_18** | 0.39847 | 0.78358 | 0.61108 | 0.91369 |
| **D\_4\_\_Erysipelotrichaceae_unclassified** | -0.94765 | 1.9043 | 0.61873 | 0.91369 |
| **D\_5\_\_Lachnoclostridium_unclassified** | -0.26616 | 0.53628 | 0.61968 | 0.91369 |
| **uncultured_bacterium_207** | 1.4393 | 2.9224 | 0.62237 | 0.91369 |
| **uncultured_bacterium_224** | -1.1179 | 2.3056 | 0.62777 | 0.91369 |
| **unidentified** | 0.69046 | 1.4262 | 0.6283 | 0.91369 |
| **uncultured_bacterium_183** | 1.2018 | 2.4887 | 0.62915 | 0.91369 |
| **Escherichia_coli** | 0.5689 | 1.179 | 0.62945 | 0.91369 |
| **human_gut_metagenome_10** | 0.39929 | 0.84224 | 0.63544 | 0.91369 |
| **uncultured_bacterium_9** | 0.17055 | 0.36353 | 0.63897 | 0.91369 |
| **Delftia_tsuruhatensis** | -1.1757 | 2.5161 | 0.64031 | 0.91369 |
| **unidentified_11** | 1.375 | 2.9462 | 0.6407 | 0.91369 |
| **D\_4\_\_Ruminococcaceae_unclassified_3** | -0.85946 | 1.8462 | 0.64156 | 0.91369 |
| **Lactobacillus_murinus** | -1.0869 | 2.3384 | 0.64207 | 0.91369 |
| **uncultured_bacterium_103** | 1.3455 | 2.9056 | 0.64331 | 0.91369 |
| **gut_metagenome_15** | 0.65645 | 1.4255 | 0.64516 | 0.91369 |
| **human_gut_metagenome_9** | -0.78795 | 1.7198 | 0.64683 | 0.91369 |
| **uncultured_Ruminococcaceae_bacterium** | 0.59449 | 1.3071 | 0.64924 | 0.91369 |
| **D\_4\_\_Ruminococcaceae_unclassified_5** | 1.3231 | 2.9132 | 0.64971 | 0.91369 |
| **uncultured_organism_73** | -0.59021 | 1.314 | 0.65332 | 0.91369 |
| **uncultured_organism_24** | 1.0053 | 2.2387 | 0.65338 | 0.91369 |
| **uncultured_organism_60** | 1.2987 | 2.8935 | 0.65355 | 0.91369 |
| **uncultured_bacterium_205** | 1.1146 | 2.4943 | 0.65498 | 0.91369 |
| **uncultured_bacterium_44** | 1.0779 | 2.4236 | 0.6565 | 0.91369 |
| **gut_metagenome_23** | -1.0095 | 2.2768 | 0.65749 | 0.91369 |
| **uncultured_bacterium_64** | -0.30777 | 0.70623 | 0.66299 | 0.91817 |
| **uncultured_bacterium_181** | 0.75994 | 1.7593 | 0.66577 | 0.91885 |
| **Intestinimonas_butyriciproducens** | -1.2067 | 2.8192 | 0.66863 | 0.91965 |
| **D\_5\_\_Ruminococcaceae_UCG_014_unclassified_2** | -1.22 | 2.925 | 0.6766 | 0.9212 |
| **Eubacterium_ramulus** | -0.35041 | 0.85121 | 0.68058 | 0.9212 |
| **uncultured_organism_16** | -0.26074 | 0.63503 | 0.68136 | 0.9212 |
| **uncultured_Clostridium_sp_** | 0.84713 | 2.0643 | 0.68154 | 0.9212 |
| **uncultured_bacterium_38** | -0.48183 | 1.181 | 0.6833 | 0.9212 |
| **uncultured_organism_19** | -0.30639 | 0.75145 | 0.68347 | 0.9212 |
| **D\_4\_\_Lachnospiraceae_unclassified** | 0.20795 | 0.54284 | 0.70166 | 0.94256 |
| **D\_4\_\_Lachnospiraceae_unclassified_1** | 0.24638 | 0.67184 | 0.71382 | 0.95571 |
| **uncultured_bacterium_30** | 0.84069 | 2.328 | 0.71801 | 0.95814 |
| **uncultured_bacterium_6** | -0.40067 | 1.1296 | 0.72281 | 0.96136 |
| **uncultured_bacterium_157** | -0.68611 | 1.9521 | 0.72523 | 0.96141 |
| **uncultured_bacterium_192** | 0.59232 | 1.7182 | 0.73029 | 0.96494 |
| **uncultured_bacterium_77** | 0.63902 | 1.8891 | 0.73516 | 0.9682 |
| **uncultured_bacterium_123** | -0.48054 | 1.4985 | 0.74845 | 0.97518 |
| **gut_metagenome_27** | -0.71753 | 2.2923 | 0.75427 | 0.97518 |
| **Lactococcus_lactis** | 0.91427 | 2.9455 | 0.75626 | 0.97518 |
| **metagenome_6** | 0.52522 | 1.7085 | 0.75852 | 0.97518 |
| **Tissierellia_bacterium_S7_1_4** | -0.6403 | 2.0847 | 0.75874 | 0.97518 |
| **gut_metagenome_38** | -0.34092 | 1.1157 | 0.75994 | 0.97518 |
| **uncultured_bacterium_177** | 0.86094 | 2.9057 | 0.767 | 0.97518 |
| **uncultured_bacterium_60** | -0.50135 | 1.7382 | 0.77302 | 0.97518 |
| **uncultured_bacterium_111** | -0.80085 | 2.8799 | 0.78095 | 0.97518 |
| **uncultured_bacterium_246** | -0.77155 | 2.8396 | 0.78584 | 0.97518 |
| **uncultured_organism_51** | -0.23942 | 0.89534 | 0.78915 | 0.97518 |
| **uncultured_bacterium_46** | 0.25136 | 0.94021 | 0.78921 | 0.97518 |
| **uncultured_organism_43** | -0.53933 | 2.0647 | 0.79393 | 0.97518 |
| **D\_4\_\_Ruminococcaceae_unclassified** | 0.344 | 1.33 | 0.79591 | 0.97518 |
| **uncultured_bacterium_36** | -0.47044 | 1.8212 | 0.79617 | 0.97518 |
| **Dubosiella_newyorkensis** | 0.24716 | 1.0025 | 0.80525 | 0.97518 |
| **uncultured_bacterium_63** | 0.69865 | 2.8898 | 0.80896 | 0.97518 |
| **uncultured_organism_65** | 0.6999 | 2.9145 | 0.81022 | 0.97518 |
| **uncultured_bacterium_130** | 0.40384 | 1.6871 | 0.81082 | 0.97518 |
| **gut_metagenome_1** | -0.22593 | 0.94404 | 0.81086 | 0.97518 |
| **uncultured_organism_85** | -0.67305 | 2.8399 | 0.81266 | 0.97518 |
| **uncultured_bacterium_239** | -0.5903 | 2.5341 | 0.81581 | 0.97518 |
| **gut_metagenome_4** | -0.48236 | 2.0827 | 0.81685 | 0.97518 |
| **uncultured_organism_33** | 0.48907 | 2.1261 | 0.81807 | 0.97518 |
| **uncultured_bacterium_23** | 0.26668 | 1.1931 | 0.82313 | 0.97518 |
| **Pseudomonas_poae** | 0.17622 | 0.80349 | 0.8264 | 0.97518 |
| **D\_4\_\_Ruminococcaceae_unclassified_1** | -0.34113 | 1.5683 | 0.82781 | 0.97518 |
| **uncultured_spirochete_1** | 0.32491 | 1.4987 | 0.82837 | 0.97518 |
| **uncultured_bacterium_137** | 0.25316 | 1.1722 | 0.82901 | 0.97518 |
| **uncultured_bacterium_190** | -0.33533 | 1.5701 | 0.83089 | 0.97518 |
| **D\_5\_\_Lachnospiraceae_NK4A136_group_unclassified** | -0.38942 | 1.8257 | 0.83109 | 0.97518 |
| **uncultured_bacterium_135** | 0.27033 | 1.2764 | 0.83227 | 0.97518 |
| **uncultured_bacterium_59** | -0.24705 | 1.1741 | 0.83335 | 0.97518 |
| **uncultured_prokaryote** | 0.20234 | 0.98222 | 0.83679 | 0.97518 |
| **uncultured_bacterium_182** | 0.31639 | 1.5619 | 0.83947 | 0.97518 |
| **Alistipes_obesi** | 0.11934 | 0.5932 | 0.84056 | 0.97518 |
| **gut_metagenome** | 0.09039 | 0.45998 | 0.84421 | 0.97518 |
| **Azospirillum_sp__47_25** | 0.56635 | 2.8998 | 0.84515 | 0.97518 |
| **uncultured_bacterium_17** | 0.22779 | 1.1692 | 0.84553 | 0.97518 |
| **uncultured_bacterium_105** | 0.34941 | 1.808 | 0.84675 | 0.97518 |
| **uncultured_bacterium_20** | -0.33948 | 1.7698 | 0.84789 | 0.97518 |
| **uncultured_Blautia_sp__1** | 0.24145 | 1.2737 | 0.84965 | 0.97518 |
| **metagenome_16** | 0.24898 | 1.3222 | 0.85063 | 0.97518 |
| **uncultured_organism_18** | 0.16509 | 0.88194 | 0.85151 | 0.97518 |
| **uncultured_organism_72** | 0.36385 | 1.9466 | 0.85173 | 0.97518 |
| **uncultured_organism_37** | -0.15119 | 0.80911 | 0.85177 | 0.97518 |
| **uncultured_bacterium_201** | -0.29575 | 1.6529 | 0.85799 | 0.97623 |
| **uncultured_bacterium_83** | -0.27791 | 1.621 | 0.86387 | 0.97623 |
| **uncultured_bacterium_284** | -0.5054 | 2.9494 | 0.86394 | 0.97623 |
| **uncultured_bacterium_57** | -0.2252 | 1.3425 | 0.86678 | 0.97623 |
| **uncultured_bacterium_106** | 0.36923 | 2.2148 | 0.8676 | 0.97623 |
| **D\_5\_\_Ruminococcaceae_UCG_002_unclassified** | 0.29001 | 1.742 | 0.86778 | 0.97623 |
| **uncultured_bacterium_48** | 0.18734 | 1.1608 | 0.87179 | 0.97623 |
| **Bacteroides_ovatus_V975_1** | 0.16248 | 1.0364 | 0.87543 | 0.97623 |
| **gut_metagenome_12** | -0.30444 | 1.9946 | 0.87869 | 0.97623 |
| **uncultured_bacterium_34** | -0.10781 | 0.71476 | 0.8801 | 0.97623 |
| **uncultured_bacterium_220** | 0.42629 | 2.9309 | 0.88436 | 0.97623 |
| **gut_metagenome_8** | 0.20166 | 1.4752 | 0.89127 | 0.97623 |
| **D\_4\_\_Lachnospiraceae_unclassified_10** | 0.39422 | 2.9158 | 0.89246 | 0.97623 |
| **Bifidobacterium_longum_subsp__longum** | -0.12576 | 0.93555 | 0.89307 | 0.97623 |
| **D\_5\_\_Bacteroides_unclassified** | 0.10073 | 0.75208 | 0.89345 | 0.97623 |
| **uncultured_organism_87** | -0.20743 | 1.5604 | 0.89425 | 0.97623 |
| **D\_5\_\_Roseburia_unclassified** | -0.38386 | 2.9163 | 0.89528 | 0.97623 |
| **metagenome_22** | -0.3301 | 2.5325 | 0.89629 | 0.97623 |
| **D\_5\_\_Bacteroides_unclassified_4** | 0.24757 | 1.9553 | 0.89924 | 0.97681 |
| **uncultured_bacterium_41** | 0.19353 | 1.6135 | 0.90452 | 0.97879 |
| **uncultured_organism_3** | 0.079325 | 0.73396 | 0.91393 | 0.97879 |
| **uncultured_bacterium_253** | -0.31492 | 2.9226 | 0.91419 | 0.97879 |
| **D\_5\_\_Bacteroides_unclassified_1** | -0.15643 | 1.513 | 0.91765 | 0.97879 |
| **D\_4\_\_Lachnospiraceae_unclassified_8** | -0.13111 | 1.2915 | 0.91914 | 0.97879 |
| **D\_5\_\_uncultured_unclassified** | 0.086169 | 0.87677 | 0.92171 | 0.97879 |
| **D\_4\_\_Lachnospiraceae_unclassified_5** | -0.25607 | 2.6155 | 0.92201 | 0.97879 |
| **uncultured_bacterium_128** | -0.22951 | 2.3916 | 0.92355 | 0.97879 |
| **uncultured_bacterium_278** | 0.27883 | 2.9433 | 0.92452 | 0.97879 |
| **gut_metagenome_13** | 0.13549 | 1.4469 | 0.9254 | 0.97879 |
| **uncultured_bacterium_11** | 0.069318 | 0.76481 | 0.92778 | 0.97879 |
| **metagenome** | 0.23838 | 2.8873 | 0.9342 | 0.98298 |
| **uncultured_bacterium_161** | 0.23032 | 2.9089 | 0.93689 | 0.98325 |
| **uncultured_bacterium_76** | -0.10001 | 1.4825 | 0.94621 | 0.98355 |
| **uncultured_organism_28** | -0.035677 | 0.53121 | 0.94645 | 0.98355 |
| **Eubacterium_ramulus_1** | -0.1167 | 1.8195 | 0.94886 | 0.98355 |
| **human_gut_metagenome_8** | 0.061326 | 0.97319 | 0.94975 | 0.98355 |
| **uncultured_Clostridium_sp__3** | 0.12995 | 2.1156 | 0.95102 | 0.98355 |
| **Prevotella_disiens_JCM_6334_=_ATCC_29426** | -0.16789 | 2.9068 | 0.95394 | 0.98355 |
| **uncultured_organism_39** | 0.05116 | 0.8919 | 0.95426 | 0.98355 |
| **uncultured_organism_6** | -0.075264 | 1.4352 | 0.95818 | 0.98506 |
| **Clostridium_phoceensis** | 0.034248 | 0.77214 | 0.96462 | 0.98819 |
| **uncultured_bacterium** | 0.063381 | 1.5463 | 0.9673 | 0.98819 |
| **uncultured_bacterium_218** | -0.11484 | 2.9145 | 0.96857 | 0.98819 |
| **uncultured_bacterium_280** | 0.080306 | 2.8405 | 0.97745 | 0.99472 |
| **uncultured_organism_52** | -0.049862 | 2.2372 | 0.98222 | 0.99706 |
| **uncultured_bacterium_50** | -0.015403 | 0.86425 | 0.98578 | 0.9975 |
| **D\_4\_\_Ruminococcaceae_unclassified_7** | -0.027749 | 2.1124 | 0.98952 | 0.9975 |
| **Lactobacillus_reuteri** | 0.036338 | 2.921 | 0.99007 | 0.9975 |
| **uncultured_bacterium_26** | 0.0029581 | 0.37945 | 0.99378 | 0.99874 |
| **uncultured_bacterium_165** | 0.0054846 | 1.7164 | 0.99745 | 0.99993 |
| **gut_metagenome_5** | 0 | 0 | 1 | 1 |

**Supplementary Table 5.** Deseq2 Analysis of Asthmatic Subjects.

|  | **log2FC** | **lfcSE** | **Pvalues** | **FDR** |
| --- | --- | --- | --- | --- |
| **Escherichia_coli** | 26.649 | 3.4483 | 1.0908E-14 | 4.3959E-12 |
| **uncultured_organism_30** | 21.245 | 2.9243 | 3.731E-13 | 7.5179E-11 |
| **unidentified** | 25.487 | 3.7242 | 7.7325E-12 | 1.0387E-09 |
| **uncultured_Alistipes_sp_** | 12.128 | 1.8144 | 2.3264E-11 | 2.3439E-09 |
| **uncultured_organism_6** | 23.93 | 3.7403 | 1.5751E-10 | 1.2696E-08 |
| **Bacteroides_dorei** | 24.249 | 3.8697 | 3.6932E-10 | 2.4358E-08 |
| **uncultured_organism_40** | 21.458 | 3.4359 | 4.2309E-10 | 2.4358E-08 |
| **D\_5\_\_Parabacteroides_unclassified** | 12.697 | 2.0818 | 1.0684E-09 | 5.3819E-08 |
| **uncultured_bacterium** | 24.963 | 4.1307 | 1.5095E-09 | 6.7593E-08 |
| **uncultured_organism_32** | 21.667 | 3.7831 | 1.0197E-08 | 4.1094E-07 |
| **uncultured_bacterium_76** | 21.022 | 3.9012 | 7.1018E-08 | 2.6018E-06 |
| **uncultured_bacterium_41** | 22.386 | 4.2111 | 1.0607E-07 | 3.5621E-06 |
| **gut_metagenome_15** | 20.936 | 3.9587 | 1.2324E-07 | 3.8203E-06 |
| **uncultured_bacterium_20** | 24.108 | 4.5785 | 1.3981E-07 | 4.0246E-06 |
| **human_gut_metagenome_6** | 20.313 | 3.8718 | 1.5504E-07 | 4.1653E-06 |
| **uncultured_bacterium_61** | 18.293 | 3.7932 | 1.4182E-06 | 3.5721E-05 |
| **uncultured_organism_34** | 21.147 | 4.5122 | 2.7769E-06 | 6.391E-05 |
| **gut_metagenome_7** | 21.755 | 4.6514 | 2.9089E-06 | 6.391E-05 |
| **human_gut_metagenome** | 24.198 | 5.1817 | 3.0131E-06 | 6.391E-05 |
| **uncultured_bacterium_34** | 10.15 | 2.1787 | 3.1794E-06 | 6.4065E-05 |
| **Bacteroides_ovatus_V975_2** | 21.706 | 4.82 | 6.6903E-06 | 0.00012839 |
| **uncultured_bacterium_94** | -21.068 | 4.7767 | 1.0312E-05 | 0.00018889 |
| **uncultured_organism_20** | -27.916 | 6.3508 | 1.1047E-05 | 0.0001899 |
| **metagenome_8** | 19.87 | 4.5256 | 1.1309E-05 | 0.0001899 |
| **uncultured_Veillonellaceae_bacterium** | 23.428 | 5.3824 | 1.345E-05 | 0.00021681 |
| **uncultured_bacterium_58** | -25.193 | 5.8814 | 1.8397E-05 | 0.00028515 |
| **gut_metagenome_19** | -26.151 | 6.154 | 2.1438E-05 | 0.00031999 |
| **uncultured_bacterium_36** | 20.883 | 5.0137 | 3.1112E-05 | 0.00044779 |
| **uncultured_bacterium_75** | 20.928 | 5.1127 | 4.2507E-05 | 0.00059069 |
| **uncultured_bacterium_37** | 24.082 | 5.9071 | 4.5668E-05 | 0.00061348 |
| **unidentified_7** | -25.345 | 6.2454 | 4.9442E-05 | 0.00064274 |
| **uncultured_bacterium_77** | 20.387 | 5.0375 | 5.1871E-05 | 0.00065325 |
| **gut_metagenome_4** | 21.082 | 5.4039 | 9.5672E-05 | 0.0011684 |
| **uncultured_bacterium_31** | 22.371 | 5.9089 | 0.00015308 | 0.0018145 |
| **uncultured_organism_24** | 22.801 | 6.0383 | 0.00015938 | 0.0018351 |
| **uncultured_Roseburia_sp_** | 17.671 | 4.7393 | 0.00019259 | 0.0021076 |
| **uncultured_organism_43** | 21.004 | 5.635 | 0.0001935 | 0.0021076 |
| **gut_metagenome_26** | 19.339 | 5.2942 | 0.00025939 | 0.0027509 |
| **Clostridium_sp__K4410_MGS_306** | 16.4 | 4.5124 | 0.00027856 | 0.0028785 |
| **uncultured_bacterium_9** | 3.0107 | 0.84921 | 0.00039221 | 0.0039516 |
| **D\_4\_\_Lachnospiraceae_unclassified_4** | 20.086 | 5.7259 | 0.00045174 | 0.0044402 |
| **D\_4\_\_Lachnospiraceae_unclassified_2** | 17.756 | 5.1304 | 0.00053834 | 0.0051655 |
| **uncultured_bacterium_21** | 10.344 | 3.0336 | 0.00064968 | 0.0060889 |
| **metagenome** | 23.529 | 6.9796 | 0.00074878 | 0.0066919 |
| **uncultured_bacterium_13** | 24.582 | 7.2946 | 0.00075205 | 0.0066919 |
| **uncultured_organism_13** | 24.823 | 7.3755 | 0.00076384 | 0.0066919 |
| **uncultured_organism_52** | 20.772 | 6.1827 | 0.00078056 | 0.0066929 |
| **gut_metagenome_27** | 20.14 | 6.0105 | 0.00080576 | 0.006765 |
| **metagenome_1** | 23.545 | 7.1115 | 0.0009302 | 0.0076504 |
| **uncultured_bacterium_44** | 20.843 | 6.3421 | 0.0010143 | 0.0081751 |
| **Ruminococcus_sp__UNK_MGS_30** | 20.548 | 6.2777 | 0.0010633 | 0.0084018 |
| **uncultured_bacterium_98** | 20.02 | 6.1936 | 0.0012274 | 0.0095124 |
| **uncultured_bacterium_63** | 21.715 | 6.7364 | 0.0012661 | 0.0096273 |
| **uncultured_bacterium_49** | -11.567 | 3.601 | 0.0013174 | 0.0096777 |
| **uncultured_organism_35** | -22.49 | 7.003 | 0.0013208 | 0.0096777 |
| **uncultured_bacterium_33** | -23.489 | 7.3956 | 0.0014925 | 0.010599 |
| **uncultured_organism_31** | -23.037 | 7.2559 | 0.0014991 | 0.010599 |
| **uncultured_bacterium_106** | 17.994 | 5.7519 | 0.0017577 | 0.012213 |
| **uncultured_bacterium_12** | 20.82 | 6.743 | 0.0020175 | 0.01378 |
| **gut_metagenome_12** | 17.124 | 5.5856 | 0.0021717 | 0.014587 |
| **D\_5\_\_Streptococcus_unclassified** | 19.568 | 6.4825 | 0.0025392 | 0.016775 |
| **gut_metagenome_17** | -21.771 | 7.2551 | 0.0026928 | 0.017503 |
| **unidentified_4** | 19.493 | 6.5501 | 0.0029208 | 0.018684 |
| **uncultured_organism_47** | 16.786 | 5.6557 | 0.0029981 | 0.018878 |
| **uncultured_bacterium_147** | 20.499 | 6.9195 | 0.0030519 | 0.018922 |
| **uncultured_bacterium_89** | 21.748 | 7.3756 | 0.0031918 | 0.019489 |
| **uncultured_bacterium_110** | 19.713 | 6.7189 | 0.0033459 | 0.020125 |
| **uncultured_bacterium_90** | 7.7604 | 2.6749 | 0.003717 | 0.022029 |
| **uncultured_bacterium_1** | 2.1013 | 0.73705 | 0.0043588 | 0.025458 |
| **uncultured_rumen_bacterium_2** | 20.142 | 7.3759 | 0.0063192 | 0.036119 |
| **uncultured_organism_2** | 4.7578 | 1.7438 | 0.0063633 | 0.036119 |
| **D\_5\_\_Prevotella_9_unclassified** | 20.064 | 7.3759 | 0.0065245 | 0.036519 |
| **uncultured_organism_49** | 20.006 | 7.3759 | 0.0066817 | 0.036887 |
| **uncultured_organism_69** | 19.905 | 7.3684 | 0.0069055 | 0.037607 |
| **Pseudomonas_poae** | 5.3314 | 1.9831 | 0.0071806 | 0.038584 |
| **uncultured_organism_38** | -19.006 | 7.1307 | 0.0076889 | 0.040771 |
| **uncultured_organism_50** | 18.401 | 6.9774 | 0.0083603 | 0.043756 |
| **uncultured_bacterium_40** | 5.0981 | 1.9561 | 0.0091537 | 0.047294 |
| **metagenome_5** | -7.6302 | 2.951 | 0.0097203 | 0.049586 |
| **gut_metagenome_28** | -12.995 | 5.1303 | 0.011307 | 0.05696 |
| **uncultured_spirochete** | -8.9364 | 3.6692 | 0.01487 | 0.073982 |
| **gut_metagenome_38** | 7.1343 | 2.9426 | 0.015331 | 0.075344 |
| **uncultured_bacterium_24** | 2.2904 | 0.95982 | 0.01702 | 0.082641 |
| **uncultured_organism_53** | 8.3208 | 3.5339 | 0.018545 | 0.08897 |
| **metagenome_11** | 6.7647 | 3.0344 | 0.025792 | 0.12228 |
| **uncultured_bacterium_107** | 7.1871 | 3.2576 | 0.027365 | 0.12823 |
| **uncultured_bacterium_85** | 8.6009 | 3.9232 | 0.028356 | 0.13088 |
| **bacterium_enrichment_culture_clone_LA92** | 6.471 | 2.9558 | 0.02858 | 0.13088 |
| **uncultured_organism_16** | 3.4142 | 1.5666 | 0.029307 | 0.13271 |
| **metagenome_7** | 7.7656 | 3.6804 | 0.034862 | 0.1561 |
| **uncultured_bacterium_11** | 3.8969 | 1.8846 | 0.038661 | 0.17121 |
| **uncultured_bacterium_109** | -8.444 | 4.2837 | 0.048702 | 0.21334 |
| **gut_metagenome_13** | 7.482 | 3.8319 | 0.050875 | 0.22046 |
| **gut_metagenome_25** | -11.838 | 6.1938 | 0.05598 | 0.24 |
| **gut_metagenome_10** | -6.4689 | 3.402 | 0.05724 | 0.24282 |
| **uncultured_bacterium_4** | 3.1699 | 1.7326 | 0.067312 | 0.28221 |
| **D\_5\_\_Blautia_unclassified** | 1.8707 | 1.0248 | 0.067926 | 0.28221 |
| **uncultured_bacterium_159** | -7.332 | 4.0424 | 0.069715 | 0.28669 |
| **D\_4\_\_Ruminococcaceae_unclassified_1** | 7.2941 | 4.0403 | 0.07102 | 0.2891 |
| **uncultured_bacterium_27** | -2.6073 | 1.4504 | 0.072229 | 0.29108 |
| **uncultured_Firmicutes_bacterium** | 6.1762 | 3.5933 | 0.085649 | 0.34175 |
| **uncultured_organism_58** | 6.2705 | 3.7423 | 0.093823 | 0.37069 |
| **Bacteroides_dorei_1** | 4.6723 | 2.8008 | 0.095277 | 0.37248 |
| **uncultured_bacterium_180** | 5.9102 | 3.5519 | 0.096123 | 0.37248 |
| **uncultured_organism_5** | 2.3941 | 1.5072 | 0.1122 | 0.43062 |
| **uncultured_bacterium_144** | 5.6861 | 3.6603 | 0.12031 | 0.45741 |
| **Bacteroides_thetaiotaomicron** | 1.9407 | 1.2632 | 0.12446 | 0.4615 |
| **gut_metagenome_23** | -8.3741 | 5.458 | 0.12496 | 0.4615 |
| **metagenome_9** | -10.42 | 6.8024 | 0.12556 | 0.4615 |
| **uncultured_Ruminococcaceae_bacterium** | -4.5878 | 2.9982 | 0.12597 | 0.4615 |
| **uncultured_organism_87** | 6.1671 | 4.052 | 0.12801 | 0.46477 |
| **uncultured_Ruminococcaceae_bacterium_1** | -6.6318 | 4.3937 | 0.13121 | 0.4721 |
| **Faecalibacterium_prausnitzii** | -6.6288 | 4.4191 | 0.13361 | 0.47399 |
| **gut_metagenome_35** | -6.8587 | 4.578 | 0.13408 | 0.47399 |
| **uncultured_organism_7** | 4.6444 | 3.129 | 0.13772 | 0.48263 |
| **uncultured_bacterium_130** | 6.5887 | 4.464 | 0.13996 | 0.48623 |
| **human_gut_metagenome_9** | 6.6647 | 4.541 | 0.14219 | 0.48978 |
| **uncultured_rumen_bacterium** | -3.7339 | 2.5642 | 0.14534 | 0.49639 |
| **metagenome_15** | -5.8271 | 4.04 | 0.1492 | 0.50528 |
| **D\_3\_\_Clostridiales_unclassified_1** | -5.5095 | 3.8349 | 0.15081 | 0.50646 |
| **D\_5\_\_Christensenellaceae_R_7_group_unclassified** | -4.7875 | 3.3648 | 0.1548 | 0.51556 |
| **uncultured_bacterium_73** | -10.339 | 7.361 | 0.16015 | 0.52902 |
| **uncultured_bacterium_201** | 5.9673 | 4.3301 | 0.16817 | 0.55101 |
| **gut_metagenome_20** | -9.9432 | 7.2539 | 0.17045 | 0.55398 |
| **D\_4\_\_Lachnospiraceae_unclassified_8** | 4.6263 | 3.3987 | 0.17345 | 0.55919 |
| **metagenome_4** | 2.9419 | 2.197 | 0.18056 | 0.57627 |
| **uncultured_organism_19** | 2.5084 | 1.8778 | 0.1816 | 0.57627 |
| **uncultured_bacterium_35** | -6.675 | 5.0244 | 0.184 | 0.57718 |
| **uncultured_organism_22** | -9.8085 | 7.3956 | 0.18476 | 0.57718 |
| **uncultured_organism_63** | -9.519 | 7.254 | 0.18943 | 0.58724 |
| **uncultured_bacterium_127** | 6.988 | 5.358 | 0.19216 | 0.58957 |
| **uncultured_organism_59** | -9.4143 | 7.254 | 0.19435 | 0.58957 |
| **uncultured_organism_10** | 1.0972 | 0.84747 | 0.19543 | 0.58957 |
| **D\_5\_\_Oxalobacter_unclassified** | 4.819 | 3.7272 | 0.19604 | 0.58957 |
| **uncultured_organism_84** | -7.5524 | 6.0689 | 0.21334 | 0.63397 |
| **bacterium_YE57** | -5.0458 | 4.0654 | 0.21455 | 0.63397 |
| **unidentified_10** | 6.5361 | 5.3345 | 0.22048 | 0.63397 |
| **uncultured_bacterium_197** | 5.6553 | 4.6302 | 0.22193 | 0.63397 |
| **human_gut_metagenome_3** | -8.8516 | 7.2541 | 0.22238 | 0.63397 |
| **uncultured_bacterium_70** | -8.8463 | 7.2542 | 0.22266 | 0.63397 |
| **uncultured_Eubacterium_sp_** | 1.7564 | 1.4425 | 0.22336 | 0.63397 |
| **unidentified_2** | 2.955 | 2.4376 | 0.2254 | 0.63397 |
| **uncultured_bacterium_16** | 1.47 | 1.2141 | 0.22598 | 0.63397 |
| **uncultured_bacterium_165** | 5.4718 | 4.5246 | 0.22653 | 0.63397 |
| **uncultured_organism** | 0.99666 | 0.8311 | 0.23045 | 0.63632 |
| **gut_metagenome_11** | -3.0443 | 2.5433 | 0.2313 | 0.63632 |
| **gut_metagenome_49** | 5.1415 | 4.3027 | 0.23211 | 0.63632 |
| **Streptococcus_salivarius_subsp__thermophilus** | 3.5047 | 2.9936 | 0.2417 | 0.65813 |
| **Parabacteroides_johnsonii_CL02T12C29** | -8.4113 | 7.2543 | 0.24626 | 0.66183 |
| **uncultured_bacterium_164** | -8.4099 | 7.2543 | 0.24634 | 0.66183 |
| **uncultured_bacterium_54** | 3.3239 | 2.891 | 0.25025 | 0.6658 |
| **uncultured_rumen_bacterium_1** | -5.3054 | 4.6424 | 0.25311 | 0.6658 |
| **uncultured_bacterium_177** | -6.5289 | 5.7311 | 0.25462 | 0.6658 |
| **uncultured_organism_26** | 0.65753 | 0.57913 | 0.25622 | 0.6658 |
| **uncultured_organism_1** | 1.0277 | 0.90617 | 0.25675 | 0.6658 |
| **uncultured_bacterium_215** | 5.1774 | 4.5746 | 0.25773 | 0.6658 |
| **D\_4\_\_Clostridiales_vadinBB60_group_unclassified_2** | -7.4983 | 6.6919 | 0.2625 | 0.67296 |
| **uncultured_bacterium_132** | 5.9276 | 5.305 | 0.26384 | 0.67296 |
| **D\_5\_\_uncultured_unclassified_5** | 5.7733 | 5.2213 | 0.26885 | 0.68025 |
| **gut_metagenome** | -1.2502 | 1.1336 | 0.27008 | 0.68025 |
| **D\_4\_\_Ruminococcaceae_unclassified_5** | -6.8422 | 6.2605 | 0.27443 | 0.68692 |
| **D\_4\_\_Ruminococcaceae_unclassified_6** | -6.6458 | 6.1421 | 0.27925 | 0.69334 |
| **gut_metagenome_52** | 4.8693 | 4.5113 | 0.28043 | 0.69334 |
| **D\_4\_\_Lachnospiraceae_unclassified** | 1.4395 | 1.3586 | 0.28937 | 0.71107 |
| **uncultured_organism_55** | 5.8843 | 5.5968 | 0.29309 | 0.71585 |
| **uncultured_bacterium_149** | -3.7114 | 3.5561 | 0.29664 | 0.72016 |
| **uncultured_organism_17** | -1.9233 | 1.8514 | 0.29887 | 0.72122 |
| **D\_4\_\_Lachnospiraceae_unclassified_5** | 5.7459 | 5.5626 | 0.30162 | 0.72354 |
| **uncultured_Clostridium_sp__1** | 5.6866 | 5.5581 | 0.30625 | 0.72578 |
| **gut_metagenome_60** | -6.3525 | 6.2114 | 0.30644 | 0.72578 |
| **D\_5\_\_Alistipes_unclassified_3** | 6.5976 | 6.4713 | 0.30796 | 0.72578 |
| **uncultured_organism_85** | 6.2287 | 6.2049 | 0.31546 | 0.73511 |
| **Delftia_tsuruhatensis** | 5.6985 | 5.678 | 0.31557 | 0.73511 |
| **uncultured_bacterium_128** | -5.056 | 5.0898 | 0.32054 | 0.74046 |
| **D\_5\_\_uncultured_unclassified** | 2.1839 | 2.2031 | 0.32154 | 0.74046 |
| **uncultured_organism_18** | -2.0942 | 2.1711 | 0.33476 | 0.76485 |
| **uncultured_bacterium_69** | 7.0977 | 7.3762 | 0.33593 | 0.76485 |
| **uncultured_organism_12** | -2.8462 | 2.9764 | 0.33894 | 0.76737 |
| **uncultured_organism_11** | 0.89695 | 0.94387 | 0.34196 | 0.76989 |
| **uncultured_bacterium_80** | -5.5503 | 5.8831 | 0.34547 | 0.77346 |
| **uncultured_organism_33** | -4.8775 | 5.2449 | 0.3524 | 0.78463 |
| **D\_4\_\_Ruminococcaceae_unclassified** | -2.9964 | 3.3002 | 0.36391 | 0.80249 |
| **uncultured_organism_81** | 6.6152 | 7.2935 | 0.36441 | 0.80249 |
| **Tissierellia_bacterium_S7_1_4** | 4.8842 | 5.4308 | 0.36847 | 0.80271 |
| **uncultured_bacterium_187** | -6.2956 | 7.0005 | 0.36849 | 0.80271 |
| **D\_4\_\_Erysipelotrichaceae_unclassified** | 4.2975 | 4.8035 | 0.37096 | 0.80376 |
| **Dubosiella_newyorkensis** | -2.1811 | 2.4875 | 0.38057 | 0.80722 |
| **uncultured_organism_15** | 2.8676 | 3.2777 | 0.38164 | 0.80722 |
| **uncultured_bacterium_84** | -6.4638 | 7.3956 | 0.38211 | 0.80722 |
| **Slackia_sp__NATTS** | 4.6402 | 5.3093 | 0.38212 | 0.80722 |
| **Prevotella_disiens_JCM_6334_=_ATCC_29426** | 6.414 | 7.3766 | 0.38457 | 0.80722 |
| **unidentified_6** | -4.4905 | 5.1645 | 0.38458 | 0.80722 |
| **Bacteroides_ovatus_V975_1** | -2.2247 | 2.5833 | 0.38914 | 0.80868 |
| **uncultured_bacterium_173** | 4.6096 | 5.4039 | 0.39365 | 0.80868 |
| **D\_4\_\_Ruminococcaceae_unclassified_7** | 4.7591 | 5.5981 | 0.39525 | 0.80868 |
| **uncultured_Clostridium_sp_** | 4.6058 | 5.4188 | 0.39534 | 0.80868 |
| **uncultured_bacterium_28** | 2.1577 | 2.5464 | 0.39679 | 0.80868 |
| **Clostridium_phoceensis** | 1.6301 | 1.9278 | 0.39779 | 0.80868 |
| **uncultured_bacterium_157** | 4.3236 | 5.1299 | 0.39933 | 0.80868 |
| **bacterium_YE57_1** | 5.9635 | 7.1161 | 0.40202 | 0.80985 |
| **uncultured_bacterium_8** | -2.4013 | 2.8771 | 0.40392 | 0.80985 |
| **uncultured_organism_23** | -1.8206 | 2.1922 | 0.40624 | 0.81047 |
| **metagenome_25** | 4.9952 | 6.1112 | 0.41371 | 0.81964 |
| **Lactobacillus_murinus** | 4.8907 | 5.9988 | 0.41491 | 0.81964 |
| **uncultured_bacterium_55** | -2.6126 | 3.2395 | 0.41997 | 0.8256 |
| **uncultured_bacterium_125** | 3.1365 | 3.9411 | 0.42612 | 0.82897 |
| **uncultured_bacterium_38** | 2.3701 | 2.9875 | 0.42758 | 0.82897 |
| **uncultured_bacterium_166** | -3.5653 | 4.4967 | 0.42786 | 0.82897 |
| **uncultured_archaeon** | -2.6381 | 3.3741 | 0.43429 | 0.83741 |
| **D\_5\_\_uncultured_unclassified_1** | 4.5698 | 5.9239 | 0.44045 | 0.84294 |
| **uncultured_bacterium_68** | -4.8153 | 6.2542 | 0.44134 | 0.84294 |
| **uncultured_bacterium_111** | 5.6529 | 7.3774 | 0.44353 | 0.84312 |
| **gut_metagenome_24** | -1.8916 | 2.485 | 0.44653 | 0.84314 |
| **uncultured_organism_104** | -5.444 | 7.2001 | 0.44959 | 0.84314 |
| **uncultured_bacterium_134** | 5.5616 | 7.3775 | 0.45093 | 0.84314 |
| **uncultured_bacterium_184** | 4.7815 | 6.3578 | 0.45201 | 0.84314 |
| **uncultured_bacterium_81** | 2.0545 | 2.7438 | 0.454 | 0.84314 |
| **uncultured_rumen_bacterium_3** | -4.4224 | 5.9392 | 0.4565 | 0.8439 |
| **D\_5\_\_GCA_900066225_unclassified** | -5.2577 | 7.1827 | 0.46417 | 0.85416 |
| **D\_4\_\_Ruminococcaceae_unclassified_3** | -3.2488 | 4.5972 | 0.47976 | 0.87338 |
| **D\_4\_\_Lachnospiraceae_unclassified_10** | 5.2095 | 7.378 | 0.48014 | 0.87338 |
| **uncultured_bacterium_131** | 5.1979 | 7.378 | 0.48112 | 0.87338 |
| **uncultured_organism_41** | 1.7647 | 2.5485 | 0.48866 | 0.87595 |
| **uncultured_bacterium_200** | 3.8157 | 5.6571 | 0.5 | 0.87595 |
| **uncultured_bacterium_51** | -2.007 | 2.9837 | 0.50117 | 0.87595 |
| **uncultured_bacterium_161** | 4.356 | 6.508 | 0.50328 | 0.87595 |
| **gut_metagenome_44** | 4.8949 | 7.3786 | 0.50709 | 0.87595 |
| **D\_5\_\_Ruminiclostridium_6_unclassified** | 3.0597 | 4.6192 | 0.50772 | 0.87595 |
| **Dorea_formicigenerans_ATCC_27755** | 0.51592 | 0.78305 | 0.50999 | 0.87595 |
| **D\_4\_\_Lachnospiraceae_unclassified_14** | 4.829 | 7.3788 | 0.51283 | 0.87595 |
| **uncultured_organism_65** | -4.7075 | 7.2422 | 0.51568 | 0.87595 |
| **uncultured_organism_60** | -3.9262 | 6.081 | 0.5185 | 0.87595 |
| **uncultured_bacterium_137** | -1.8488 | 2.8736 | 0.51998 | 0.87595 |
| **uncultured_bacterium_193** | 4.7245 | 7.379 | 0.522 | 0.87595 |
| **uncultured_bacterium_23** | 1.7901 | 2.8249 | 0.52629 | 0.87595 |
| **uncultured_organism_64** | -3.7073 | 5.8531 | 0.52649 | 0.87595 |
| **uncultured_bacterium_232** | -2.6663 | 4.2485 | 0.53028 | 0.87595 |
| **uncultured_Blautia_sp__1** | -1.9621 | 3.1329 | 0.53113 | 0.87595 |
| **Bifidobacterium_bifidum** | -2.7684 | 4.4327 | 0.53227 | 0.87595 |
| **uncultured_organism_21** | -2.4534 | 3.9657 | 0.53614 | 0.87595 |
| **uncultured_organism_9** | -1.1134 | 1.8058 | 0.53752 | 0.87595 |
| **uncultured_organism_29** | 1.0734 | 1.7478 | 0.53912 | 0.87595 |
| **uncultured_bacterium_48** | -1.7717 | 2.9025 | 0.5416 | 0.87595 |
| **D\_2\_\_Clostridia_unclassified** | -3.9227 | 6.4483 | 0.54297 | 0.87595 |
| **uncultured_bacterium_269** | -4.4003 | 7.2652 | 0.54473 | 0.87595 |
| **uncultured_organism_36** | 2.7247 | 4.4999 | 0.54484 | 0.87595 |
| **D\_4\_\_Ruminococcaceae_unclassified_10** | 4.4427 | 7.3798 | 0.54717 | 0.87595 |
| **uncultured_bacterium_181** | -2.6951 | 4.4801 | 0.54746 | 0.87595 |
| **uncultured_bacterium_66** | 1.1406 | 1.9082 | 0.55002 | 0.87595 |
| **uncultured_bacterium_59** | -1.7237 | 2.9012 | 0.55242 | 0.87595 |
| **uncultured_bacterium_2** | 0.7242 | 1.2198 | 0.55271 | 0.87595 |
| **metagenome_14** | 4.2956 | 7.3803 | 0.56054 | 0.87595 |
| **uncultured_bacterium_6** | -1.6497 | 2.8421 | 0.56161 | 0.87595 |
| **Lactobacillus_reuteri** | -4.1857 | 7.257 | 0.56408 | 0.87595 |
| **D\_5\_\_Ruminococcaceae_UCG_010_unclassified_1** | 4.255 | 7.3804 | 0.56426 | 0.87595 |
| **uncultured_bacterium_220** | -4.1426 | 7.2491 | 0.56768 | 0.87595 |
| **gut_metagenome_46** | 3.5772 | 6.2605 | 0.56774 | 0.87595 |
| **uncultured_bacterium_280** | 4.2098 | 7.3806 | 0.56842 | 0.87595 |
| **uncultured_bacterium_205** | 3.8344 | 6.7492 | 0.56995 | 0.87595 |
| **Bifidobacterium_longum_subsp__longum_1** | -1.4345 | 2.5412 | 0.57242 | 0.87595 |
| **uncultured_bacterium_170** | 4.165 | 7.3807 | 0.57254 | 0.87595 |
| **Azospirillum_sp__47_25** | -3.9351 | 6.9838 | 0.57312 | 0.87595 |
| **uncultured_organism_95** | 4.0997 | 7.381 | 0.57859 | 0.87595 |
| **D\_4\_\_Lachnospiraceae_unclassified_12** | 3.4092 | 6.1514 | 0.57943 | 0.87595 |
| **D\_5\_\_Blautia_unclassified_3** | 3.7967 | 6.8933 | 0.58178 | 0.87595 |
| **unidentified_3** | 1.8755 | 3.4155 | 0.58293 | 0.87595 |
| **uncultured_bacterium_25** | 3.0184 | 5.5375 | 0.5857 | 0.87595 |
| **uncultured_Clostridium_sp__2** | -2.2289 | 4.0996 | 0.58665 | 0.87595 |
| **Bacteroides_fragilis** | 2.7314 | 5.0348 | 0.58748 | 0.87595 |
| **uncultured_Clostridium_sp__3** | -2.8938 | 5.3665 | 0.58973 | 0.87595 |
| **uncultured_bacterium_101** | -2.4578 | 4.5821 | 0.59169 | 0.87595 |
| **uncultured_bacterium_218** | 3.939 | 7.3816 | 0.5936 | 0.87595 |
| **gut_metagenome_9** | 0.98147 | 1.8409 | 0.59393 | 0.87595 |
| **uncultured_bacterium_207** | 3.9182 | 7.3817 | 0.59556 | 0.87595 |
| **uncultured_bacterium_3** | -0.78866 | 1.531 | 0.60647 | 0.88864 |
| **uncultured_bacterium_246** | -3.3672 | 6.5799 | 0.60883 | 0.88864 |
| **gut_metagenome_1** | -1.2106 | 2.3787 | 0.6108 | 0.88864 |
| **uncultured_bacterium_210** | 3.6632 | 7.3829 | 0.61977 | 0.89845 |
| **Eubacterium_ramulus_1** | 2.2805 | 4.6387 | 0.62298 | 0.89986 |
| **Clostridium_sp__CAG:306** | 3.6042 | 7.3832 | 0.62543 | 0.90018 |
| **uncultured_bacterium_18** | 0.94439 | 1.9721 | 0.63202 | 0.90642 |
| **uncultured_bacterium_235** | 3.1737 | 6.7949 | 0.64045 | 0.91525 |
| **D\_4\_\_Lachnospiraceae_unclassified_1** | 0.78355 | 1.693 | 0.64349 | 0.91635 |
| **uncultured_bacterium_142** | 3.3395 | 7.3848 | 0.65112 | 0.92311 |
| **uncultured_organism_46** | -2.0163 | 4.4822 | 0.65282 | 0.92311 |
| **uncultured_bacterium_87** | 1.6947 | 3.802 | 0.65577 | 0.92404 |
| **gut_metagenome_3** | -1.1129 | 2.5155 | 0.65818 | 0.92421 |
| **Bacteroides_ovatus_V975_3** | -2.1822 | 4.9916 | 0.66198 | 0.92624 |
| **uncultured_organism_51** | 0.9631 | 2.2218 | 0.66466 | 0.92624 |
| **uncultured_organism_25** | -1.6764 | 3.9001 | 0.66732 | 0.92624 |
| **uncultured_bacterium_32** | -1.2513 | 2.9512 | 0.67157 | 0.92624 |
| **uncultured_bacterium_156** | -3.0722 | 7.2828 | 0.67313 | 0.92624 |
| **uncultured_bacterium_115** | -2.0168 | 4.7854 | 0.67342 | 0.92624 |
| **uncultured_bacterium_182** | -1.6512 | 3.9699 | 0.67746 | 0.9285 |
| **Lactobacillus_gasseri** | -0.62566 | 1.5153 | 0.67967 | 0.9285 |
| **uncultured_organism_88** | 3.0208 | 7.387 | 0.68259 | 0.92933 |
| **human_gut_metagenome_4** | -1.8747 | 4.6349 | 0.68587 | 0.93065 |
| **uncultured_bacterium_183** | -1.9648 | 5.1072 | 0.70044 | 0.94503 |
| **Ruminococcus_sp__DJF_VR70k1** | 1.9441 | 5.0783 | 0.70186 | 0.94503 |
| **uncultured_bacterium_105** | -1.7617 | 4.6286 | 0.7035 | 0.94503 |
| **Lactococcus_lactis** | -2.6561 | 7.2925 | 0.71569 | 0.95111 |
| **uncultured_bacterium_67** | -1.478 | 4.067 | 0.7163 | 0.95111 |
| **uncultured_bacterium_45** | 0.48107 | 1.3315 | 0.71787 | 0.95111 |
| **uncultured_bacterium_57** | -1.2023 | 3.4078 | 0.72424 | 0.95111 |
| **uncultured_organism_61** | -1.1359 | 3.2326 | 0.72531 | 0.95111 |
| **uncultured_Ruminococcus_sp_** | 0.91479 | 2.6421 | 0.72916 | 0.95111 |
| **uncultured_organism_3** | 0.62688 | 1.8132 | 0.72954 | 0.95111 |
| **Streptococcus_pneumoniae** | 2.5458 | 7.3909 | 0.7305 | 0.95111 |
| **D\_5\_\_Lachnospiraceae_NK4A136_group_unclassified** | 1.545 | 4.5144 | 0.73217 | 0.95111 |
| **D\_5\_\_Methylobacterium_unclassified** | 2.4929 | 7.3912 | 0.73591 | 0.95111 |
| **Bacteroides_ovatus_V975** | -1.1553 | 3.4304 | 0.73628 | 0.95111 |
| **uncultured_Blautia_sp_** | 0.26427 | 0.78609 | 0.73673 | 0.95111 |
| **uncultured_organism_72** | 1.6166 | 4.8508 | 0.73894 | 0.95111 |
| **metagenome_18** | 2.4425 | 7.3916 | 0.74106 | 0.95111 |
| **uncultured_rumen_bacterium_5** | -1.058 | 3.2911 | 0.74785 | 0.9539 |
| **metagenome_16** | -1.0722 | 3.337 | 0.74797 | 0.9539 |
| **uncultured_bacterium_50** | 0.68573 | 2.1842 | 0.75356 | 0.95793 |
| **uncultured_bacterium_26** | -0.2908 | 0.93881 | 0.75675 | 0.95793 |
| **uncultured_bacterium_15** | -1.519 | 4.9358 | 0.75826 | 0.95793 |
| **D\_5\_\_Ruminiclostridium_9_unclassified** | -1.1507 | 3.979 | 0.77243 | 0.96599 |
| **uncultured_bacterium_239** | 2.1328 | 7.3933 | 0.77298 | 0.96599 |
| **D\_4\_\_Lachnospiraceae_unclassified_3** | -1.0913 | 3.7928 | 0.77355 | 0.96599 |
| **uncultured_bacterium_253** | 1.895 | 6.6061 | 0.77423 | 0.96599 |
| **uncultured_bacterium_17** | -0.79358 | 2.8713 | 0.78225 | 0.9692 |
| **uncultured_Clostridiales_bacterium** | 1.2604 | 4.5635 | 0.7824 | 0.9692 |
| **uncultured_organism_45** | 0.52575 | 1.933 | 0.78563 | 0.9692 |
| **D\_3\_\_Clostridiales_unclassified_5** | -1.566 | 5.7996 | 0.78714 | 0.9692 |
| **uncultured_Thermoanaerobacterales_bacterium_1** | 1.7607 | 6.574 | 0.78883 | 0.9692 |
| **gut_metagenome_58** | -1.7628 | 6.9998 | 0.80117 | 0.98002 |
| **Alistipes_obesi** | -0.35312 | 1.4944 | 0.8132 | 0.98002 |
| **uncultured_bacterium_190** | 0.89196 | 3.9074 | 0.81944 | 0.98002 |
| **gut_metagenome_22** | -0.86582 | 3.8258 | 0.82096 | 0.98002 |
| **metagenome_13** | -0.91897 | 4.1615 | 0.82523 | 0.98002 |
| **D\_5\_\_Alistipes_unclassified_2** | -0.71303 | 3.2326 | 0.82542 | 0.98002 |
| **uncultured_organism_73** | -0.72853 | 3.3557 | 0.82813 | 0.98002 |
| **uncultured_organism_48** | 0.64329 | 2.9728 | 0.82868 | 0.98002 |
| **D\_5\_\_Bacteroides_unclassified_1** | 0.82513 | 3.842 | 0.82995 | 0.98002 |
| **uncultured_bacterium_284** | 1.5465 | 7.3954 | 0.83436 | 0.98002 |
| **Eubacterium_ramulus** | 0.44606 | 2.1553 | 0.83604 | 0.98002 |
| **uncultured_bacterium_64** | 0.33772 | 1.7793 | 0.84947 | 0.98002 |
| **Alistipes_indistinctus_YIT_12060** | -0.72844 | 3.8589 | 0.85027 | 0.98002 |
| **uncultured_bacterium_172** | -1.0919 | 5.7878 | 0.85037 | 0.98002 |
| **human_gut_metagenome_2** | -1.0375 | 5.5346 | 0.8513 | 0.98002 |
| **metagenome_6** | 0.81903 | 4.3871 | 0.8519 | 0.98002 |
| **uncultured_bacterium_238** | -1.3172 | 7.2424 | 0.85568 | 0.98002 |
| **human_gut_metagenome_8** | -0.43657 | 2.4196 | 0.85681 | 0.98002 |
| **uncultured_bacterium_278** | 1.3232 | 7.3956 | 0.858 | 0.98002 |
| **uncultured_bacterium_257** | 1.3231 | 7.3956 | 0.85801 | 0.98002 |
| **metagenome_22** | -1.0728 | 6.0768 | 0.85987 | 0.98002 |
| **gut_metagenome_2** | -0.29417 | 1.7222 | 0.86437 | 0.98002 |
| **uncultured_prokaryote** | -0.42362 | 2.4882 | 0.86481 | 0.98002 |
| **D\_5\_\_Bacteroides_unclassified_4** | -0.83768 | 4.9777 | 0.86636 | 0.98002 |
| **uncultured_spirochete_1** | 0.64638 | 3.8584 | 0.86696 | 0.98002 |
| **uncultured_rumen_bacterium_4** | 0.70502 | 4.5766 | 0.87757 | 0.98002 |
| **uncultured_bacterium_123** | -0.57875 | 3.8302 | 0.8799 | 0.98002 |
| **Bifidobacterium_longum_subsp__longum** | -0.35005 | 2.3664 | 0.8824 | 0.98002 |
| **uncultured_organism_28** | -0.19512 | 1.3373 | 0.884 | 0.98002 |
| **uncultured_organism_8** | -0.48831 | 3.5602 | 0.89091 | 0.98002 |
| **D\_5\_\_Lachnoclostridium_unclassified** | -0.19919 | 1.5561 | 0.89815 | 0.98002 |
| **uncultured_bacterium_42** | 0.58087 | 4.5734 | 0.89893 | 0.98002 |
| **uncultured_bacterium_83** | 0.51245 | 4.1485 | 0.90169 | 0.98002 |
| **uncultured_bacterium_60** | -0.50841 | 4.5379 | 0.91079 | 0.98002 |
| **Intestinimonas_butyriciproducens** | 0.7049 | 6.4046 | 0.91236 | 0.98002 |
| **uncultured_bacterium_154** | 0.70488 | 6.4265 | 0.91266 | 0.98002 |
| **uncultured_bacterium_121** | -0.49733 | 4.5372 | 0.91272 | 0.98002 |
| **D\_5\_\_Roseburia_unclassified** | 0.70485 | 6.5004 | 0.91365 | 0.98002 |
| **uncultured_bacterium_39** | -0.13686 | 1.3541 | 0.9195 | 0.98002 |
| **uncultured_bacterium_198** | 0.70478 | 7.2465 | 0.92252 | 0.98002 |
| **D\_5\_\_Ruminococcaceae_UCG_014_unclassified_2** | 0.70481 | 7.2703 | 0.92277 | 0.98002 |
| **uncultured_bacterium_249** | 0.70483 | 7.3246 | 0.92334 | 0.98002 |
| **unidentified_14** | 0.70483 | 7.326 | 0.92335 | 0.98002 |
| **uncultured_bacterium_178** | 0.70493 | 7.3956 | 0.92406 | 0.98002 |
| **unidentified_11** | 0.70495 | 7.3956 | 0.92406 | 0.98002 |
| **uncultured_Bacteroides_sp_** | 0.70487 | 7.3956 | 0.92407 | 0.98002 |
| **unidentified_9** | 0.70486 | 7.3956 | 0.92407 | 0.98002 |
| **D\_5\_\_Tyzzerella_4_unclassified** | 0.70478 | 7.3956 | 0.92408 | 0.98002 |
| **uncultured_bacterium_82** | 0.7048 | 7.3956 | 0.92408 | 0.98002 |
| **uncultured_bacterium_103** | 0.70473 | 7.3956 | 0.92408 | 0.98002 |
| **human_gut_metagenome_12** | 0.7048 | 7.3956 | 0.92408 | 0.98002 |
| **D\_4\_\_Lachnospiraceae_unclassified_13** | 0.70481 | 7.3956 | 0.92408 | 0.98002 |
| **uncultured_bacterium_224** | -0.4374 | 6.1188 | 0.94301 | 0.98842 |
| **uncultured_bacterium_135** | 0.22146 | 3.1819 | 0.94451 | 0.98842 |
| **uncultured_bacterium_5** | 0.075064 | 1.1087 | 0.94602 | 0.98842 |
| **unidentified_1** | -0.13709 | 2.0532 | 0.94677 | 0.98842 |
| **uncultured_bacterium_192** | -0.29686 | 4.4482 | 0.94679 | 0.98842 |
| **uncultured_organism_39** | 0.15018 | 2.2566 | 0.94694 | 0.98842 |
| **uncultured_organism_27** | 0.17498 | 2.8768 | 0.9515 | 0.98842 |
| **uncultured_bacterium_30** | -0.36829 | 6.0714 | 0.95163 | 0.98842 |
| **uncultured_bacterium_46** | 0.11915 | 2.3483 | 0.95953 | 0.99211 |
| **D\_5\_\_Alistipes_unclassified** | -0.09013 | 2.0687 | 0.96525 | 0.99211 |
| **gut_metagenome_8** | 0.13422 | 3.5747 | 0.97005 | 0.99211 |
| **D\_5\_\_Bacteroides_unclassified** | 0.068607 | 1.8761 | 0.97083 | 0.99211 |
| **uncultured_bacterium_138** | 0.17175 | 5.4416 | 0.97482 | 0.99211 |
| **uncultured_organism_14** | -0.041555 | 1.7251 | 0.98078 | 0.99211 |
| **human_gut_metagenome_10** | -0.050393 | 2.1047 | 0.9809 | 0.99211 |
| **uncultured_bacterium_43** | -0.12313 | 5.2395 | 0.98125 | 0.99211 |
| **uncultured_bacterium_14** | 0.01538 | 0.66598 | 0.98157 | 0.99211 |
| **uncultured_organism_37** | -0.046839 | 2.0463 | 0.98174 | 0.99211 |
| **uncultured_bacterium_79** | 0.088887 | 3.9979 | 0.98226 | 0.99211 |
| **Anaerostipes_hadrus** | 0.016495 | 1.8609 | 0.99293 | 1 |
| **D\_5\_\_Ruminococcaceae_UCG_002_unclassified** | 0.0082947 | 4.4118 | 0.9985 | 1 |
| **uncultured_organism_4** | 0.0024239 | 2.6201 | 0.99926 | 1 |
| **gut_metagenome_5** | 0 | 0 | 1 | 1 |
